# Pitfalls in estimating and interpreting the contribution of ultra-rare genetic variants to the heritability of complex traits

**DOI:** 10.64898/2026.04.06.26350278

**Authors:** Huanwei Wang, Pierrick Wainschtein, Julia Sidorenko, Mulusew Fikere, Yuanxiang Zhang, Kathryn E. Kemper, Zhili Zheng, Valentin Hivert, Jian Zeng, Michael E. Goddard, Peter M. Visscher, Loic Yengo

**Affiliations:** Institute for Molecular Bioscience, University of Queensland, Brisbane, QLD, Australia; Statistical Genetics Group, Population Health Program, QIMR Berghofer, Brisbane, Queensland, Australia; Illumina Artificial Intelligence Laboratory, Illumina Inc., San Deigo, CA, USA; Purdue University, West Lafayette, IN 47907, USA; Broad Institute of MIT and Harvard, Boston, Massachusetts, USA; Faculty of Veterinary and Agricultural Science, University of Melbourne, Parkville, Victoria, Australia; Nuffield Department of Population Health, University of Oxford, Oxford, UK

## Abstract

Assessing the contribution of ultra-rare variants (minor allele frequency <0.01%) to the heritability of complex traits remains challenging due to limited understanding of potential biases. Here, we focus on singletons (that is, variants observed only once in the study sample), the most abundant class of ultra-rare variants, to showcase various confounders of heritability estimates and underline pitfalls in their interpretation. We show through theory, simulations, and analysis of 5,330,210 exome-sequenced singletons in 305,813 unrelated European-ancestry individuals in the UK Biobank that (i) population stratification induces both upward and downward biases in singleton-based heritability estimates 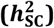, (ii) 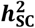 estimates capture non-additive genetic effects, and (iii) asymptotic standard errors of 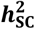 estimates from likelihood-based procedures are generally mis-calibrated when traits are not normally distributed. We further showcase these biases in real-data analyses of 22 quantitative phenotypes and report, after accounting for these pitfalls, significant 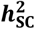 estimate for number of children (3.4%), peak expiratory flow (1.9%), red blood cell count (2.5%), white blood cell count (1.9%) and heel bone mineral density (2.4%). Overall, our study provides recommendations for robust inference of heritability from ultra-rare variants and underscores that reliable estimates for ordinal and binary traits will require far larger sample sizes and improved methods, given that confounding in these traits remains difficult to detect and correct.

## INTRODUCTION

The contribution of rare genetic variation to inter-individual trait differences has yet to be fully quantified. Recent studies in European ancestry populations have provided precise estimates of heritability explained by variants with a minor allele frequency (MAF) larger than 0.01% and have identified dozens of traits for which family-based heritability is fully accounted for by variants ascertainable by whole-genome sequencing (WGS) technologies^1^. Nevertheless, a knowledge gap remains for variants with a MAF<0.01% (hereafter referred to as ultra-rare variants), which constitute the majority of genetic variants.

Reliable heritability estimates from ultra-rare variants are currently missing from the literature because of limited sample sizes and unexpected findings from studies that have attempted to quantify their contribution. For example, Wainschtein and colleagues^1^ observed significantly negative estimates for 17 traits, which typically indicates model misspecification^2^. However, the exact misspecification at play remains unknown. Similarly, Rocheleau and colleagues^3^ imposed strong constraints for the estimation of coronary artery disease heritability from ultra-rare variants by using an EM algorithm (guaranteed to produce non-negative estimates) for more than 10,000 iterations, which also indicates numerical instability. These two examples illustrate the need for a clear and robust framework to obtain and correctly interpret heritability estimates from ultra-rare variants.

The present study addresses some of the root causes of these unexpected estimates of heritability and numerical instabilities by proposing new theory and by performing extensive simulations and real-data analyses to expose pitfalls in the estimation of heritability from ultra-rare variants. Without loss of generality, we focus on singleton variants (that is, variants observed only once in a sample) because they account for 40-50% of all ultra-rare variants,^4^ their analysis is computationally efficient to enable estimations from large samples and previous studies have suggested they may account for a substantial amount of gene expression heritability. ^5^ Overall, we show that nonsensical heritability estimates (for example, negative or larger than one) can be commonly caused by biases induced by population stratification, scale effect and non-normal phenotypic distributions and propose various sensitivity approaches and adjustments to minimize these issues.

## RESULTS

### Overview of the heritability model

We assume the additive effect *b*_*j*_ of each singleton *j* to be random and independently drawn from a distribution with mean *µ*_*b*_ and variance 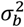. Under these assumptions, the expectation and variance of the focal trait (*y*_*i*_) of individual *i*, conditionally on all their singletons, can be expressed as a function of individual *i*’s singleton count (SC), hereafter denoted *S*_*i*_, as follows:

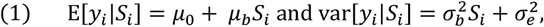

where *µ*_0_ and 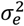 are the expectation and variance of non-singletons effects on *y*_*i*_ . The parameters of the model defined in Equation (1) can be estimated using methods of moments or likelihood-based approaches. Likelihood-based approaches require additional assumptions about the distribution of *y*_*i*_|*S*_*i*_ (that is *y*_*i*_ conditionally on *S*_*i*_) and that of *b*_*j*_. Here, we classically assume that both *y*_*i*_|*S*_*i*_ and *b*_*j*_ are independent and normally distributed, where *y*_*i*_ represents the observed phenotypes (or the underlying liability in the case of a binary trait), and therefore, use the Genome-based Restricted Maximum Likelihood (GREML)^6,7^ as our primary method for estimation throughout this manuscript. We describe a computationally optimised implementation of GREML for analysing singletons in the **METHODS** section. Our approach differs from the burden heritability regression method^8^, which assumes the same allele effect sizes within genes, and thus only captures a component of total heritability explained by variation between genes.

In theory, the contribution of singletons to the overall genetic variance of *y* depends on their potential directional effect ( *µ*_*b*_ ). It can be expected if, on average, singletons tend to be deleterious for fitness. However, in practice, these directional effects can be accounted for by modelling SC as a fixed effect in GREML analyses. Therefore, for simplicity and consistency with previous GREML studies, we focus on the non-directional component of singleton effects and hereafter refer to 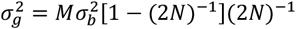 as the genetic variance attributable to all singletons, where *M* is the total number of singletons in the study of sample size *N*. Note that the term [1 − (2*N*)^−1^](2*N*)^−1^represents the sampling variance of allele count for each singleton, which is assumed to follow a Bernoulli distribution with mean (2*N*)^−1^.

Finally, we define the singleton-based heritability as 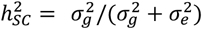 . Importantly, our model cannot distinguish the additive effect of singletons from that of epistatic interactions between all singletons carried by a given individual. Therefore, 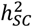 does not solely contribute to the narrow sense heritability but also to the broad sense heritability.

### Confounding due to population stratification

#### Theoretical expectations

Equation (1) implies that factors simultaneously associated with SC and affecting the variance of *y* can be confounders for 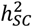 estimates 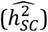. In particular, population structure can lead sub-groups in the sample to have more or less singletons than average because of genetic drift. Therefore, if the phenotypic distribution in one or more of these sub-groups also differs from that in the overall sample, then biases in 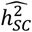 are expected. We can illustrate this point using a simplified model of population stratification (PS) in which the study sample is composed of two subgroups of individuals differing in both their average SCs and phenotypic means. If we denote *π* as the proportion of individuals in the smallest group (i.e., 0 < *π* < 0.5), Δ_*S*_ as the difference in mean SC between the smallest and the largest group, and 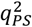 the proportion of phenotypic variance explained by PS, then the expectation of 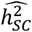 when 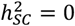 can be expressed as

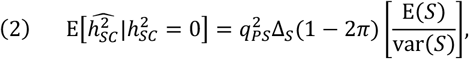

where E(*S*) and var(*S*) denote the mean and variance of SC across the entire sample. Importantly, while it has been previously reported that population stratification can bias heritability estimates[refs^9,10^], the direction of the bias is often assumed to be positive. Equation (2) shows that biases in 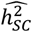 can be both positive and negative and that the sign of this bias only depends on Δ_*S*_. Moreover, this equation also predicts that the magnitude of the bias can be larger than the actual amount of variance explained by PS (that is, 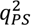) if |Δ_*S*_| > var (*S*)/[(1− 2*π*)E(*S*)]. For example, if the distribution SC is such that E(*S*) ≈ var(*S*), then 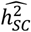 would be expected to exceed 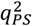 when |Δ_*S*_| > 1/(1 − 2*π*) ≈ 1 (for small values of *π* ). The proof of Equation (2) is given in the **Supplementary Note 1**. A derivation of the bias when confounding is due to a difference in phenotypic variance between subgroups is given in the **Supplementary Note 2**.

#### Performance of various PS correction methods with simulated data

We further illustrate our theoretical results using simulations based on genotypes of UK Biobank participants (**METHODS**). Beyond validating Equation (2), these simulations also aimed to assess the performances of different approaches for correcting biases in heritability estimates caused by PS. Note that previous studies have already shown that using genetic principal components can be suboptimal for correcting biases induced by localised PS in genome-wide association studies_11_ and polygenic score analyses^12^. In all simulations, 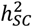 is set to be 0 and PS was simulated such that it explains 10% of phenotypic variance between two sub-groups of UK Biobank participants differing in their average SC. The smallest sub-group is chosen (as a reference group) to have a higher phenotypic mean than the other group. We considered two simulation scenarios defined by how this reference group is determined. Specifically, we used 378 birth regions (BR) of UK Biobank participants as previously defined by Abdellaoui et al. (2019)^13^ and focused on the two BRs with the lowest (Sheffield: 14.3 across 8,126 individuals) and highest (London: 23.7 across 4,135 individuals) average SC (**Supplementary Table 1**). In the first simulation scenario the reference group is defined by individuals born in *London*, while the second scenario corresponds to a reference group with individuals born in *Sheffield*.

On average across 1,000 simulation replicates, we found a significant positive 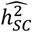 under the first scenario and a significant negative 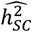 under the second scenario (**Fig. 1a**), both consistent with predictions from Equation (2). Next, we compared different approaches to correct the observed biases using various sets of covariates included in GREML analyses. These sets of covariates include genomic principal components (PC) calculated from common variants, assessment centres, SC, and k-means clusters of birthplace coordinates within the UK. While we observed a reduction of the bias for each set (and combinations thereof) of covariates fitted in the model (**Fig. 1a**), none of them seemed sufficient to completely remove it. Importantly, we found that biases could be further reduced when birthplace clusters were also fitted as random effects (**Fig. 1b**).

**Figure 1.**
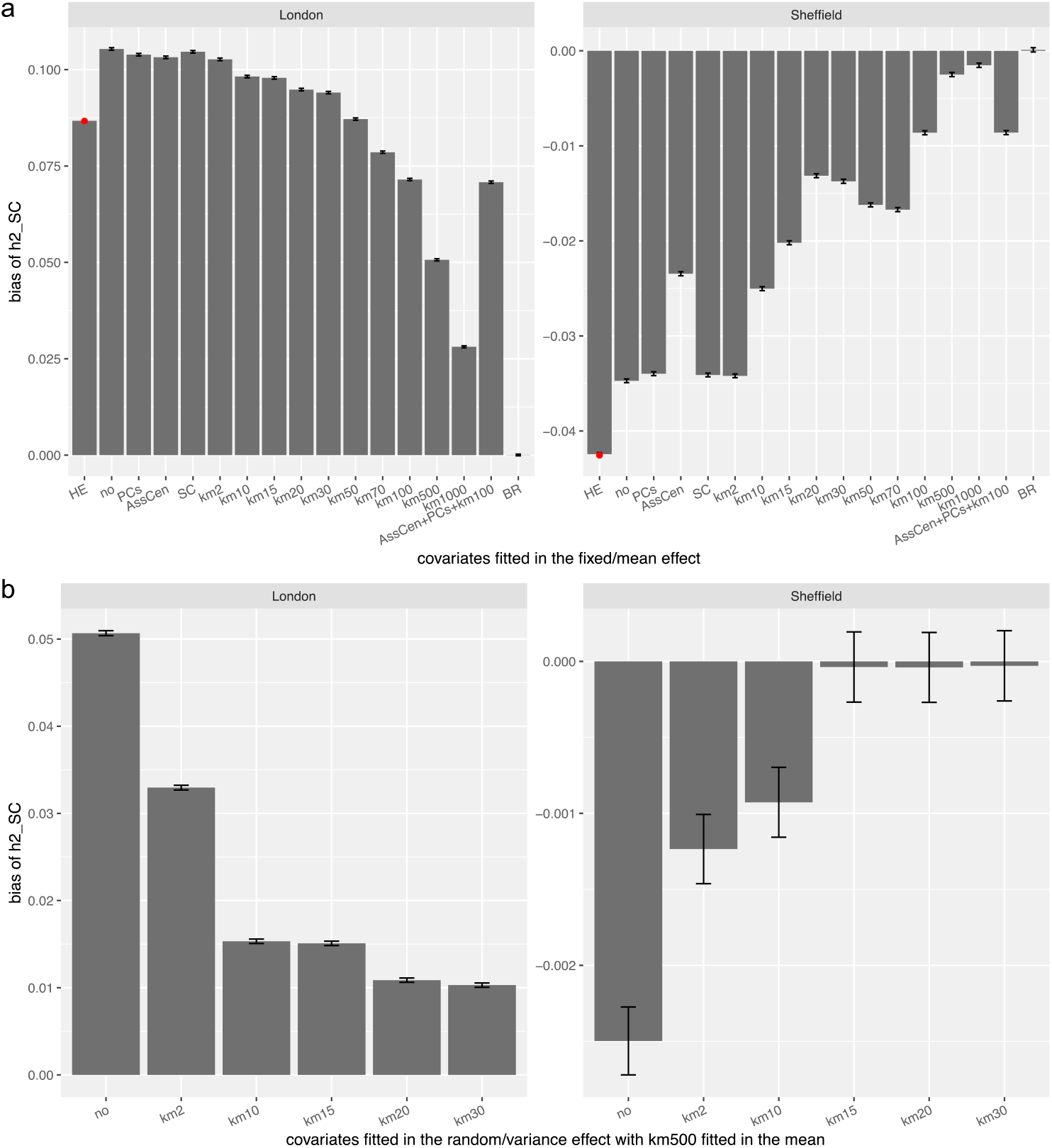
The bias of 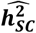 estimates in a simulation model of PS based on two birth regions (i.e. London and Sheffield). a) Haseman-Elston (HE)-regression-based estimates and GREML-based estimates with different covariates fitted as fixed effects. The red points indicate the theoretical values based on Equation (2); b) Fitting different covariates as random effects in addition to fitting km500 as fixed effect. The results were based on 1,000 simulation replications for each of two birth regions. Fitted covariates include principal components (PCs), assessment centres (AssCen), singleton count (SC), birth regions (BR), and k-means clustering based on birth coordinates (Data Fields 129 and 130) with 2, 10, 15, 20 and 30 clusters (indicated by km2, km10, km15, km20 and km30 in Pane) (**METHODS**). Error bars represent 1.96 times standard errors.

#### Effect of PS on heritability enrichment in functional genomic annotations

Given that standard methods were unable to fully correct confounding biases induced by PS, we further investigated if complementary strategies aiming at quantifying a relationship between heritability and functional genomic annotations could, at least, inform if a significant 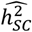 is more biologically plausible. Therefore, we assessed the effect of PS on partitioned heritability estimates across functional genomic annotations. We considered two annotations. The first annotation groups singletons into three functional classes defined as: neutral (n=1,109,609), missense (n=1,543,163) and predicted loss of function (pLOF; n=70,280), while the second annotation (hereafter denoted WES-target annotation) discriminates singletons within WES-targeted exome regions (n=2,723,052) from off-target flanking regions (n=2,607,158). These annotations are further detailed in the **METHODS** section. Enrichment in each annotation was defined as the ratio of per-SNP heritability for SNPs in the annotation relative to the overall per-SNP heritability (**METHODS**).

Before analysing simulated data, we first tested the association between these annotations and BRs to assess if biases in functional enrichment could be expected. Overall, we found a significant association between BRs and burden of pathogenic variants (Chi-square test: *P* = 2.25 × 10^−11^; **Supplementary Fig. 1**) but not between BRs and the WES-target annotation (Chi-square test: P=0.44; **Supplementary Fig. 1**). The joint-distribution of genomic annotations and BRs is reported in **Supplementary Table 2**.

On average across 1,000 simulation replicates, we found significant heritability enrichment in all annotations with magnitudes varying between 0.5-fold and 1.2-fold for GREML models without (**Fig. 2**) or with (**Supplementary Fig. 2**) fitting covariates, thus demonstrating that PS can induce spurious heritability enrichment in functional annotation.

**Figure 2.**
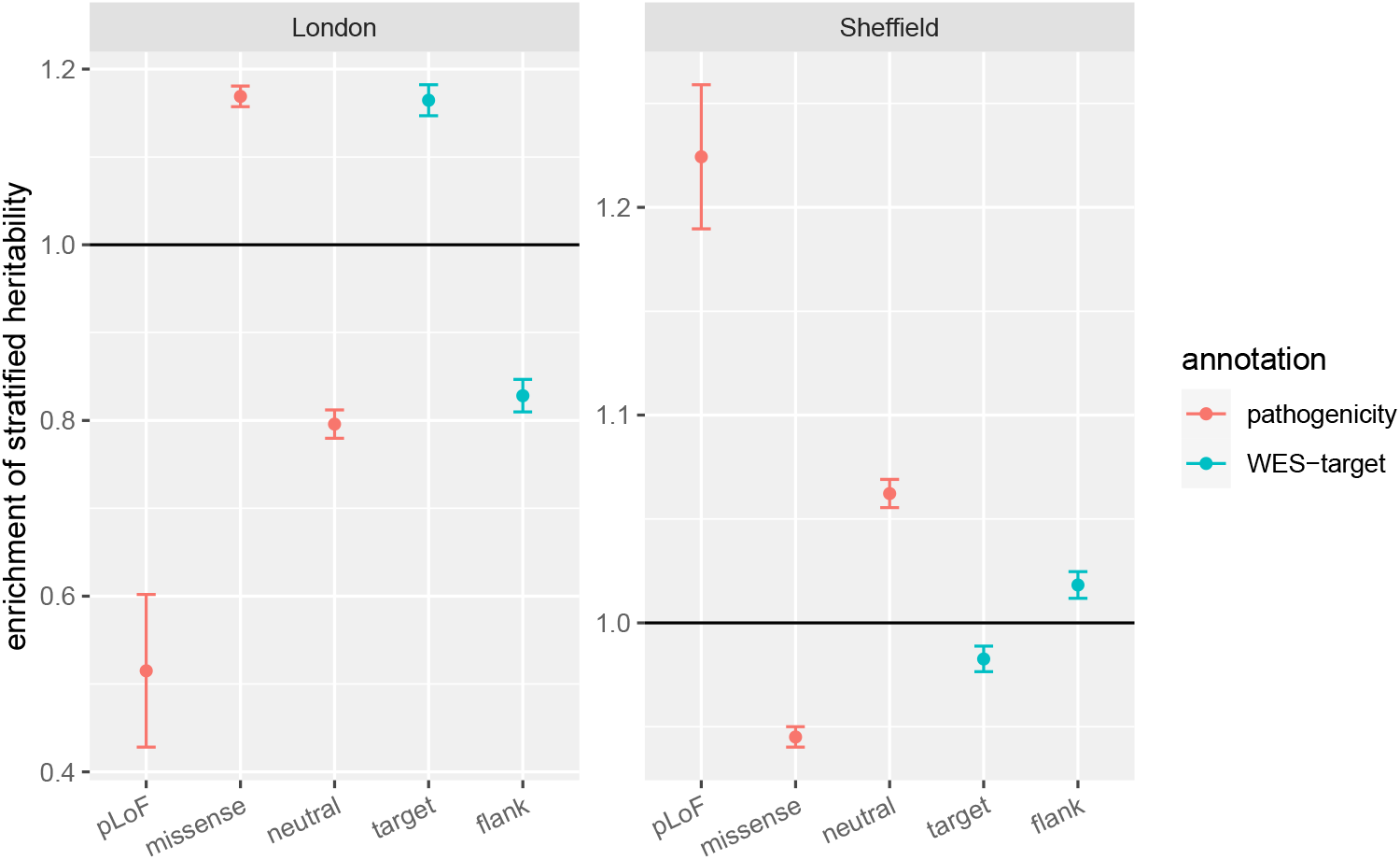
The per-SNP singleton-based heritability enrichments estimated by the GREML method for pathogenicity and WES-target annotations in the simulation model of PS. The results above were based on GREML model without covariates, and results with different covariates fitted in the GREML model can be found in **Supplementary Figure 2**. The pathogenicity functional annotations and WES target regions were downloaded from UKB (**URLs**; **METHODS**). Singletons annotated as “synonymous” or lacking annotation information were labelled as “neutral”. Error bars represent 1.96 times standard errors.

#### Impact of truncation of SC distribution

PS induces overdispersion in SC distribution because of positive covariances between singletons across the genome (**Supplementary Note 3, and Figure 3-5**). We hypothesized that outliers in the SC distribution (**METHODS**) might be caused by individuals carrying haplotypes whose genetic ancestry differs from that of the rest of the study sample. If this hypothesis is true, then excluding outliers might help reduce PS-induced biases. To test this hypothesis, we first identified a sub-group of singletons also present in the African, East-Asian and South-Asian ancestry samples from the 1000 Genomes Project^14^. We hereafter refer to those as migration singletons^15^. Overall, we found that migration singletons explain 20.3% of SC variance (**Supplementary Fig. 3**). Specifically, we found that carrying 1 migration singleton is associated with 1.8-fold increase in the probability of being a SC outlier, which supports our hypothesis.

**Figure 3.**
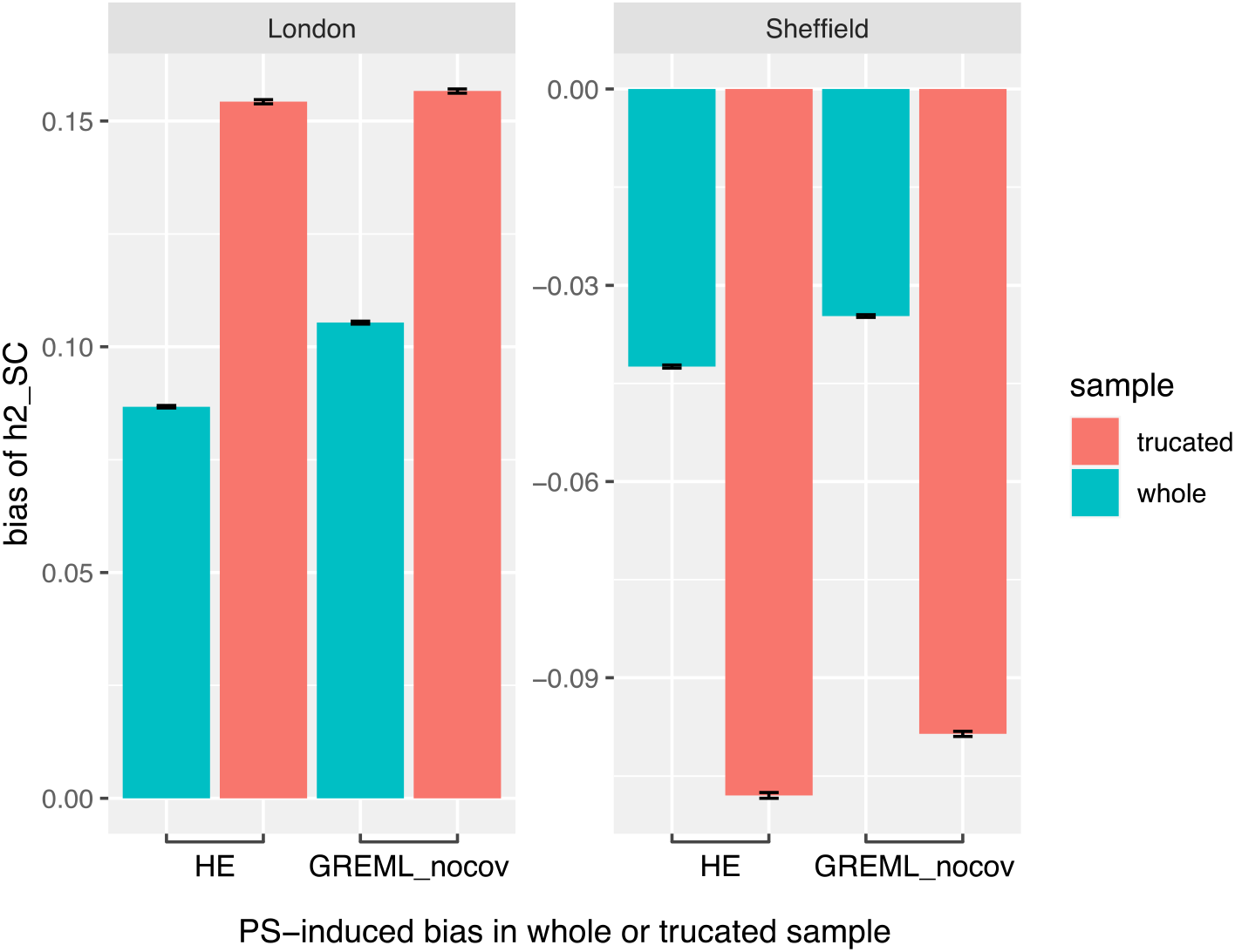
The bias of 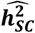 in the whole and truncated sample in the simulation model of PS. The truncated sample removed SC outliers, defined as individuals whose SC values exceed six times the median SC plus the median SC (**METHODS**). The singleton variants were re-called in the truncated sample. Error bars represent 1.96 times standard errors.

However, when assessing the impact of right-truncation of the SC distribution on 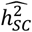, we found under our two PS simulation scenarios that excluding outliers inflates the magnitude of PS-induced bias (**Fig. 3**). Importantly, this inflation can be predicted from Equation (2) when E(*S*) and var(*S*) are recalculated in the truncated sample. Note that truncation is expected to reduce heritability in the absence of PS (**Supplementary Note 4**) when causal variants are randomly distributed among singletons. Overall, these results imply that although PS contributes to SC outliers, a simple exclusion might not resolve biases and may, in some cases, exacerbate them.

### Effect of scale transformation

We performed additional simulations to showcase scenarios, other than PS, that can yield significantly negative heritability estimates. Specifically, we show that such situations can occur when singletons tend to have directional effects (i.e., when *µ*_*b*_ ≠ 0 ) on some underlying (Gaussian) scale but the scale on which the phenotype is observed is skewed, thus inducing a mean-variance relationship. For each simulation replicate, we assumed a constant effect *b* of each singleton on an underlying Gaussian scale, which we varied from -0.01 to +0.01 (step size: 0.005) trait standard deviation (SD) per singleton. This corresponds to SC explaining less than 1.2% of variance on the underlying Gaussian scale. We then mapped phenotypic values from the underlying scale onto the observed scale of five real traits measured in the UKB: education attainment (EA), disease count (DC), height (HT), body mass index (BMI), and the number of children (NC). The mapping function is described in the **METHODS** section. All analyses include SC as fixed effect to attempt to correct for the simulated directional effect.

Over 1,000 simulation replicates, we found that negative directional effects of singletons on the liability scale can induce significantly negative 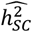 for most trait-specific scales (**Fig. 4b-e**). For the scale mimicking EA distribution in the UKB, we also observed negative 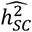 when directional effects of singletons were positive (**Fig. 4f**). We found that applying rank-based inverse normal transformation (RINT) to the phenotype can substantially reduce the magnitude of these biases for most trait-specific scales, except that of EA. Note that we use the term bias here because our definition of 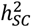 does not include directional effects. We present additional simulation scenarios for binary traits in **Supplementary Fig. 6**.

**Figure 4.**
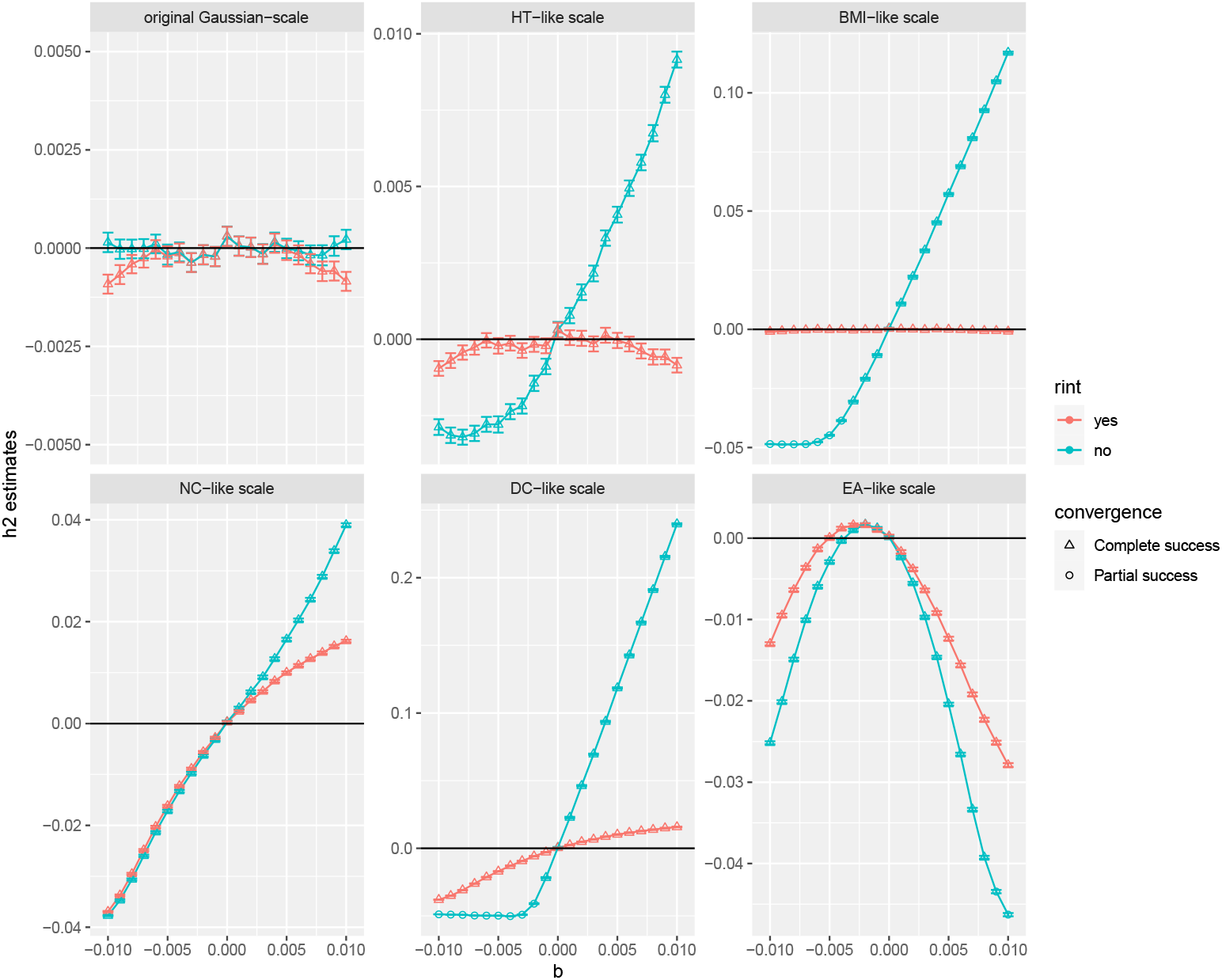
The impact of scale transformation on 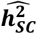 in the presence of directional effects of SC on a phenotype. We simulated the phenotype with a directional effect of SC (**METHODS**). We then mapped the simulated phenotype from the original Gaussian-scale to a transformed scale based on real distribution of each of five traits, i.e., height (HT), body mass index (BMI), the number of children (NC), disease count (DC), and education attainment (EA) (Supplementary Table 3). The 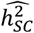 were estimated from the GREML model with SC fitted as the fixed effect to account for the directional effect. Results based on rank-based inverse transformation (rint) are shown in red, and results without transformation are shown in green. Some simulation replicates failed to converge due to the biased negative heritability estimates, which have been indicated as “partial success” in the figure. Error bars represent 1.96 times standard errors.

In summary, these simulations provide an alternative explanation for significant negative heritability estimates, highlight how directional effects of singletons can bias heritability estimates, and show that fitting SC as fixed effect only partially reduces these biases when the phenotypic distribution is skewed. Note that this scale effect can also impact the magnitudes of mean effect estimates, but not the direction (**Supplementary Fig. 7**).

### Mis-calibration of statistical significance of 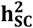 estimates

This section explores the effect of trait distribution on the statistical significance of 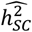. Under standard asymptotic assumptions, the standard error of maximum likelihood estimates can be approximated using the inverse of the observed Fisher information matrix. However, it has been previously reported that rare variant association tests based on variant aggregation methods such as burden tests tend to have mis-calibrated p-values for non-Gaussian traits (ref^16^). We compared asymptotic and permutation-based standard errors for different families of phenotypic distributions and found that non-Gaussian phenotypic distributions systematically yielded inflated or deflated standard errors (**Figure 5**). Therefore, we recommend using empirical standard errors, for example, obtained using permutation-based approaches.

**Figure 5.**
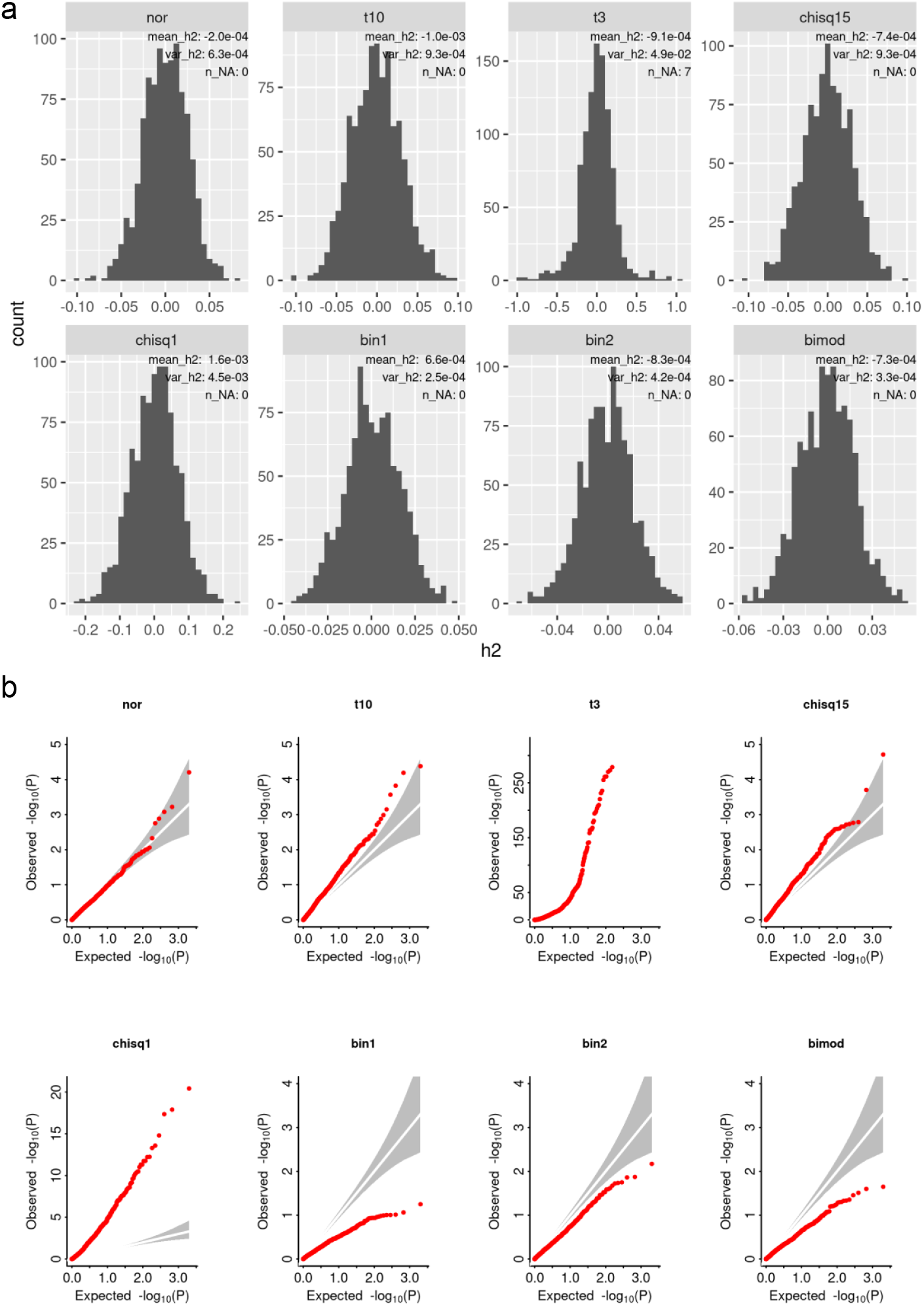
Simulation of different trait distributions on the point estimates (a) and test-statistics (b) of 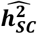 based on the GREML method. The phenotype was simulated from eight distributions: normal distribution (nor), *t*-distribution with degree of freedom (df) of 10 and 3 (t10 and t3), χ^2^ distribution with df of 15 and 1 (chisq15 and chisq1), binomial (1, 0.3) (bin1), binomial (2, 0.3) (bin2), and bimodal normal mixture distribution (0.3normal(- 3,1)+0.7normal(3,1)) (bimod). The SC was simulated from a Poisson distribution with the parameter lambda of 32. The simulation was replicated 1,000 times for a sample size of 100,000. The grey area in the QQ-plot in the panel b was the 95% confidence interval.

### Singleton-based heritability estimates for 22 UKB traits

In previous sections, we exposed various biases and pitfalls in the interpretation of 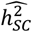 using simulated data. Here, we analyse 22 quantitative traits measured in the UKB to show that these issues are also observed in real data (**Supplementary Table 3**).

We analysed each trait using GREML, sequentially fitting covariates from the following set as fixed effects: sex, year of birth (YOB), YOB squared (YOBsq), 22 assessment centres (AssCen), 40 PCs, and 378 BRs, to account for population stratification. When fitting the full set of covariates, we identified 13 phenotypes with significant 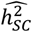 (*p*<0.05/22; **Table 1**), including six negative heritability estimates: DC, systolic blood pressure (SBP), mean time to correctly identify matches (MTCIM), birth weight (BW), EA, and speech-reception-threshold mean measurement (SRTM). Overall, no substantial changes in the of 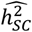 estimates were observed when fitting different sets of covariates, except for a few traits (**Figure 6**). For example, estimates of 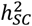 for NC decreased from 7.2% (no covariates) to 5.7% (full covariate model).

**Table 1.** 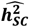 for 22 quantitative traits measured in the UK Biobank. We report the results fitting covariates as fixed effects, including sex, year of birth (YOB), YOB squared (YOBsq), 22 assessment centres (AssCen), 40 PCs, and 378 BRs (“Covariates fitted as fixed effects only” columns), and those further fitting covariates as random effects, including seven YOB groups, 22 AssCen, and 10 k-means-based birth clusters (“Covariates fitted as both fixed and random effects” columns). Permutation-based p-values were calculated as the percentage of permutations with estimates exceeding the point estimates in the original models (see **METHODS**). P values less than 0.05/22 were defined as significant and highlighted in plink. More details about phenotype definitions can be found in **Supplementary Table 3**.

| phenotype | Covariates fitted as fixed effects only |  |  |  | Covariates fitted as both fixed and random effects |  |  |  |
| --- | --- | --- | --- | --- | --- | --- | --- | --- |
| | $\widehat{h}_{sc}^2$ | Standard error | p-value | Permutation-based p-value | $\widehat{h}_{sc}^2$ | Standard error | p-value | Permutation-based p-value |
| The number of children (NC) | 0.057 | 0.0048 | 2.48E-34 | 0.00E+00 | 0.034 | 0.0045 | 2.99E-14 | 0.00E+00 |
| Disease count (DC) | -0.027 | 0.0036 | 3.90E-12 | 4.00E-04 | 0.022 | 0.0033 | 1.65E-11 | 5.32E-03 |
| Systolic blood pressure (SBP) | -0.024 | 0.0037 | 1.50E-09 | 0.00E+00 | -0.003 | 0.0034 | 3.73E-01 |  |
| Peak expiratory flow (PEF) | 0.024 | 0.0045 | 3.81E-08 | 0.00E+00 | 0.019 | 0.0041 | 3.00E-06 | 6.00E-04 |
| Mean time to correctly identify matches (MTCIM) | -0.018 | 0.0037 | 3.47E-06 | 3.00E-03 | 0.005 | 0.0034 | 1.12E-01 |  |
| Birth weight (BW) | -0.022 | 0.0048 | 9.72E-06 | 1.90E-03 | 0.007 | 0.0052 | 1.52E-01 |  |
| White blood cell count (WBC) | 0.018 | 0.0043 | 2.38E-05 | 9.00E-04 | 0.019 | 0.0045 | 1.26E-05 | 2.00E-04 |
| Education attainment (EA) | -0.016 | 0.0038 | 4.54E-05 | 0.00E+00 | 0.000 | 0.0034 | 9.02E-01 |  |
| Speech-reception-threshold estimate (mean) (SRTM) | -0.025 | 0.0061 | 9.72E-05 | 3.19E-02 | 0.006 | 0.0055 | 2.74E-01 |  |
| Red blood cell count (RBC) | 0.016 | 0.0042 | 1.10E-04 | 6.00E-04 | 0.025 | 0.0043 | 4.35E-09 | 0.00E+00 |
| Basal metabolic rate (BMR) | 0.016 | 0.0043 | 1.50E-04 | 1.60E-03 | 0.004 | 0.0041 | 3.11E-01 |  |
| Body mass index (BMI) | 0.015 | 0.0042 | 2.16E-04 | 4.60E-03 | 0.009 | 0.0042 | 2.46E-02 |  |
| Height (HT) | 0.015 | 0.0042 | 4.36E-04 | 6.00E-04 | 0.010 | 0.0040 | 1.74E-02 |  |
| Fluid intelligence score (FIS) | 0.021 | 0.0074 | 2.98E-03 | 9.00E-04 | 0.006 | 0.0070 | 3.93E-01 |  |
| logMAR (mean) (logMARM) | 0.024 | 0.0088 | 4.33E-03 | 4.99E-02 | 0.031 | 0.0086 | 3.77E-04 | 1.71E-02 |
| Forced expiratory volume (FEV) | 0.011 | 0.0044 | 8.83E-03 | 2.36E-02 | 0.008 | 0.0035 | 2.53E-02 |  |
| Heel bone mineral density (hBMD) | 0.014 | 0.0055 | 9.23E-03 | 5.52E-02 | 0.024 | 0.0056 | 1.96E-05 | 1.20E-03 |
| Hand grip strength (mean) (HGSM) | 0.009 | 0.0042 | 2.98E-02 | 3.40E-02 | 0.003 | 0.0038 | 4.09E-01 |  |
| Waist-to-hip ratio (WHR) | 0.008 | 0.0041 | 6.45E-02 | 8.13E-02 | 0.005 | 0.0040 | 2.47E-01 |  |
| Neuroticism score (NTS) | 0.006 | 0.0045 | 1.76E-01 | 1.12E-01 | 0.004 | 0.0046 | 3.56E-01 |  |
| Pulse rate (PR) | -0.007 | 0.0067 | 3.14E-01 | 3.99E-01 | -0.004 | 0.0070 | 5.77E-01 |  |
| Diastolic blood pressure (DBP) | -0.003 | 0.0041 | 4.56E-01 | 4.77E-01 | -0.001 | 0.0042 | 7.41E-01 |  |

**Figure 6.**
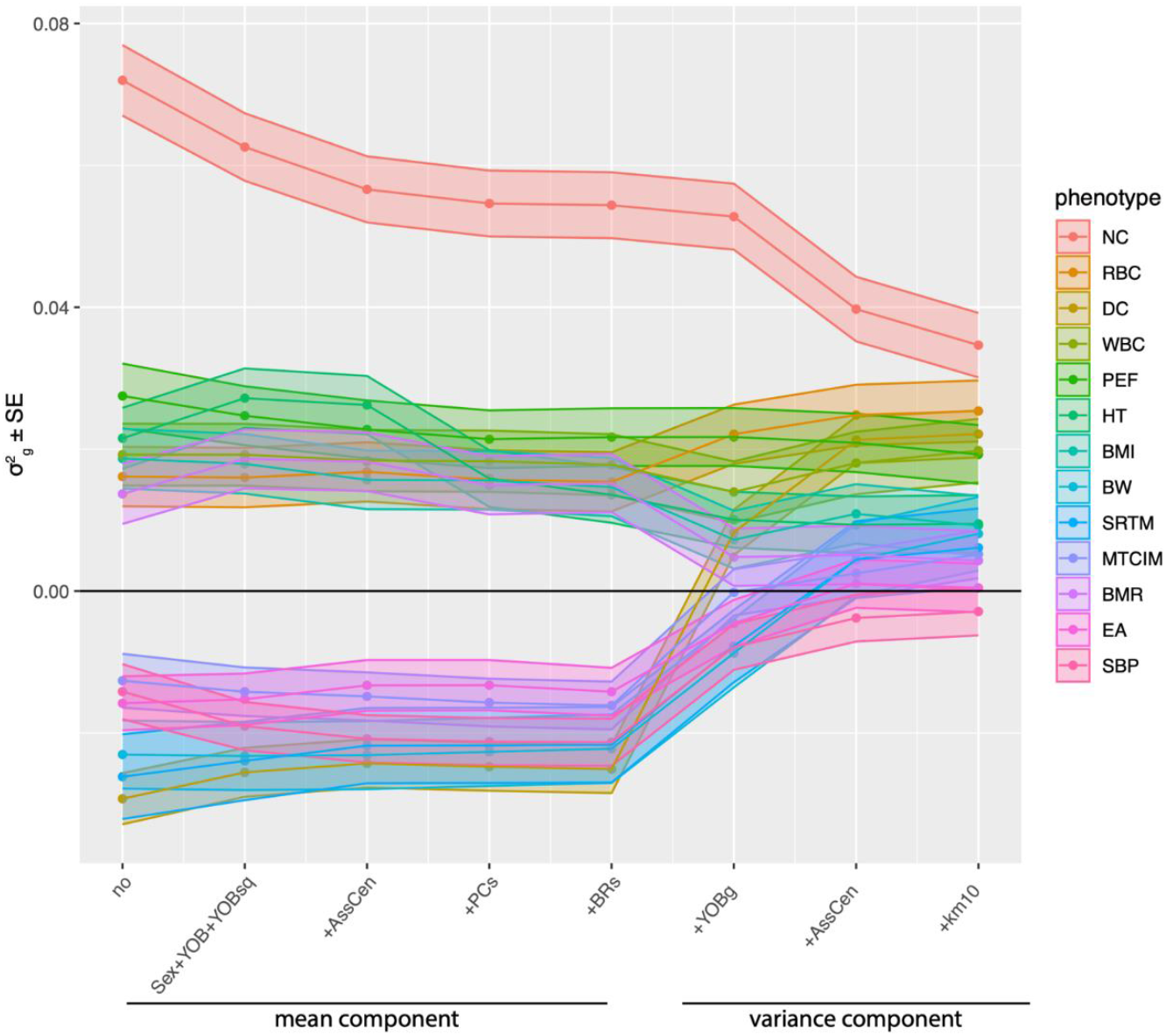
Fitting different covariates as fixed and/or random effects in the GREML models for 13 traits. We sequentially fitted covariates as fixed effects from sex, year of birth (YOB), YOB squared (YOBsq), 22 assessment centres (AssCen), top 40 PCs, and 378 BRs, and then sequentially added covariates as random effects from seven YOB groups (YOBg), assessment centres, and 10 k-means BC clusters. The genetic variance estimates 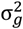 and their GREML standard errors were showed. These 13 traits were selected because they showed a statistically significant non-zero 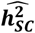 in the GREML model with covariates fitted as fixed effects only (Table 1). Confidence band represent standard errors from the GREML model.

We also re-assessed the statistical significance of 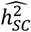 using permutations and found that 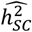 for 3 traits (including negative estimates for 2 traits) no longer passed our significance threshold, suggesting an underestimation of their sampling variance was the most likely explanation of these findings. Interestingly, we observed an overestimation of the asymptotic sampling variance of 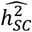 for fluid intelligence score (**Table 1**).

We then sequentially included seven year-of-birth groups (YOBg), 22 assessment centres, and 10 k-means-based birth clusters (see **METHODS**) as random effects, in addition to the full set of covariates previously fitted as fixed effects. Under these models, all six previously negative 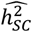 became non-significant except for DC whose heritability estimate reversed direction from a significantly negative estimate of -2.7% to a significantly positive estimate of 2.2% (**Figure 6**). This observation is consistent with the nonlinear confounding effects shown in **Fig. 1**. Furthermore, estimates of heritability for two traits no longer passed our significance threshold after applying permutation-based testing in a model including all three additional random effects. We also observed a further reduction in 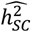 for NC, which decreased from 5.7% down to 3.4% (**Table 1**).

We found that RINT reduced the occurrence of negative estimates. Only two traits showed significantly negative 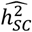 when the full set of covariates were included as fixed effects, and none remained significant when adding additional random effects (**Supplementary Table 4 and Figure 8**). Across the 22 traits, the magnitude of the 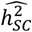 estimates were generally reduced under RINT compared to non-RINT, suggesting potential bias introduced by scale effects, as shown in our simulations (**Supplementary Figure 9**). Note that we only observed significant directional effect of SC for three traits, i.e., DC, FIS, and SBP (**Supplementary Table 5**).

Next, we assessed the effect of right-truncation of the SC distribution on our heritability estimates. Overall, we observed an increase in the magnitude of 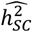 (**Supplementary Figure 10**), consistent with potential PS-induced bias shown in (**Fig. 3**).

Finally, we partitioned 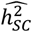 for four traits with a significant singleton-based heritability estimates (NC, peak expiratory flow (PEF), red blood cell count (RBC), and white blood cell count (WBC); **Table 1**). As in our simulations, we grouped variants based on predicted pathogenicity and their location relative to loci targeted by WES technologies. Overall, these analyses did not show a significant heritability enrichment in any of the annotations (**Supplementary Table 6**).

## DISCUSSION

In this study, we showcase various biases affecting heritability estimates from singleton variants. We developed new theory to predict the direction of biases due to uncorrected population stratification and presented simulations showing the effect of scale transformation on heritability estimates. We highlight scenarios of population stratification where singleton counts correlate with differences in phenotypic variance between subgroups in the population and show, in these scenarios, that standard approaches based on fixed-effects correction may be inefficient. Therefore, we recommend fitting covariates as random effects as a sensitivity analysis to ensure that confounding due to population stratification has been effectively corrected.

It is worth noting that including additional random effects in the analyses also makes the interpretation of heritability estimates more challenging. For example, if the phenotypic variance changes across the levels of a certain factor (e.g., year of birth), then the heritability of that trait is not uniquely defined. Nevertheless, fitting this factor as random effect would still reduce biases affecting estimates of genetic variance (**Fig. 6**) and thus improve the estimation of heritability within each level of the covariates.

Our study shows that GREML-based estimates (and their expected standard errors) from singletons can be highly sensitive to the distribution of the trait, especially when singletons tend to exhibit directional effects. Importantly, whereas rank-based transformations can reduce these biases, they cannot fully correct them. One possible solution to this problem would be to use generalized linear mixed models (such as double generalized linear model (DGLM)^17^ or hierarchical generalized linear model (HGLM)^18,19^), which allow to specify a family of distributions for the trait analysed. These classes of models have been used previously to detect genetic variants associated with trait variability^20^. However, the latter methods may still be influenced by the choice of the link function and a misspecification of the trait distribution. Future works using liability threshold models (using multiple thresholds^21^) may allow a more robust inference agnostic to the choice of distribution but not that of the link function (e.g., probit or logit). Overall, these challenges make the estimation of the ultra-rare variant heritability for binary and categorical traits much less reliable than that of quantitative traits.

Our study has a several limitations. First, we focused on singletons available through WES, whose distribution may differ from that of other singletons across the genome. Nevertheless, the biases highlighted here are expected to generalise to all singletons (and ultra-rare variants) as they mainly reflect the effect of trait distribution and that of differences in mean and variance of allele counts between groups of individuals (effect of population stratification) and across genomic loci (effect of genetic architecture). Second, our study did not address other factors such as ascertainment of study participants, which might also induce biases in heritability estimates. In fact, the known healthy- and educated-volunteer bias in the UK Biobank might have contributed to the downward biases in estimates of singleton-based heritability for disease count and educational attainment (**Table 1**).

Despite these challenges and limitations, our study and that of Wainschtein and colleagues^1^ suggest that the magnitude of singleton-based heritability remains lower than 5% (average across traits; **Table 1**) and, therefore, may not substantially contribute to the gap between pedigree-based and SNP-based heritability also known as missing heritability. Nevertheless, our study highlights the need for new statistical models to reliability quantify this remaining component of the missing heritability. Developing such methods would open new avenues for quantifying the effect of *de novo* mutations on trait variation, a long-standing question in quantitative genetics.

## METHODS

### UK Biobank Whole Exome Sequencing data

The UK Biobank (UKB) is a prospective cohort study comprising genetic and phenotypic data for approximately 500,000 individuals aged between 40 and 69 years at the time of recruitment during 2007 to 2010. Genotyping was initially performed by an array-based method followed by imputation, with data released in 2018^22^. This was succeeded by Whole Exome Sequencing (WES) and Whole Genome Sequencing of the entire cohort in 2022 (ref.^23^) and late 2023 (ref.^24^), respectively. In this study, we used the final release of WES data, provided in PLINK format (Data Field 23158) through the UKB research analysis platform (RAP). This dataset includes 469,835 individuals and 26,388,327 autosomal variants.

We selected a subset of unrelated individuals of European ancestry, previously identified using imputed genotype data by projecting the UKB PCs onto PCs of the 1000 Genome projects (1KG) and removing one of each pair of individuals with HapMap 3 SNP-based GRM > 0.05 using GCTA^25^ (**URLs**). In this study, PS was a key confounder, and we accounted for it using various covariates, including birthplace coordinates (BCs; Data Fields 129 and 130) and its derived variables such as BR and k-means clusters of BCs. We observed substantial differences in SC between individuals with and without BC information (data not showed). Therefore, we restricted our analysis to individuals with available BC data, resulting in a final subset of 305,813 individuals.

We identified 5,330,210 biallelic autosomal singleton SNPs using PLINK2^26^ (“--mac 1 --max-mac 1” option). Among these, 2,723,051 singletons were located within 197,212 autosomal target regions captured on the UKB exome panel (**URLs**), which span approximately 37.5 Mb and range in size from 1 bp to ∼20.9 kb. The pathogenicity functional annotations for these singletons were extracted from helper file “ukb23158_500k_OQFE.annotations.txt” (resource 916; see **URLs**) provided on the UKB RAP, which contains annotations for 8,187,996 autosomal variants. As the majority of these variants (7,959,762) are located within the target regions, we restricted our pathogenicity-stratified heritability estimation analysis to singletons only located within target regions. Singletons annotated as “synonymous” or lacking annotation information were grouped into a “neutral” category [ref^23^].

SCs for each individual were computed using the “--sample-counts” function implemented in PLINK2. The distribution of SC exhibited substantial overdispersion with SC mean 17.43 and SD 11.14 (**Supplementary Note 3**). To define a truncated dataset with outliers removed, we applied a threshold of six times the median SC away from the median. Mean and variance are equal under a Poisson distribution, and the median is more robust to skewness than the mean, serving as a more reliable estimation of the location parameter. In the truncated set of 294,916 individuals, we re-called the singletons, resulting in 4,887,098 singletons with SC mean 16.57 and SD 6.07.

We selected 22 quantitative traits for our analysis (**Supplementary Table 3**). We removed phenotypic outliers 5 SD away from the mean, standardized to mean 0 and variance 1 in each gender group, and conducted a second round of standardization across the whole sample. We considered various covariates as fixed and/or random effects in the models. These included basic demographic variables (sex, year of birth (YOB) and YOB square) as well as covariates accounting for PS (PCs derived from common variants, 22 assessment centres and 378 BRs). When fitting covariates as random effects, including too many random components caused GREML to fail to converge. To address that, we grouped YOB into seven categories (YOBg) and applied k-means clustering to BCs. We selected 10 clusters in real data analysis and explored different numbers of k-means BC clusters in the simulation study.

### 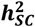 estimation by GREML model

The standard GREML approach^6,7^ to estimate SNP-based heritability fitted all variants as random effects in a linear mixed model:

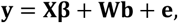

where **y** was a *N*-dimensional vector of phenotypes with *N* being the sample size, **X** was a *N* × *K* matrix of covariates as fixed effects, **β** was a *K*-dimensional vector of fixed effect sizes, **W** was a *N* × *M* matrix of standardized genotypes with *M* being the total number of variants, ***b*** was a *M*-vector of random effect sizes with 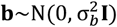, and ***e*** was a *N*-vector of residual terms with 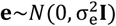. The phenotypic variance is

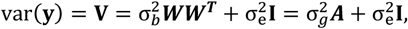

where ***A*** = ***WW***^***T***^/*M* denotes the genetic relationship matrix (GRM) and 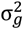 the variance of the genetic component. So, heritability was defined by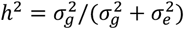.

In the case of singleton variants, the GRM was

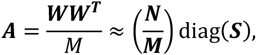

with ***S*** being a N-vector of SCs. By leveraging the fact that the GRM matrix ***A*** is diagonal, resulting in a diagonal phenotypic variance matrix ***V***, we developed a *diagGREML()* function in R (**URLs**) based on average information (AI)-REML algorithm^27^, which significantly accelerated computation.

For stratified 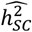 estimation, we fitted multiple GRMs 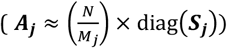 based on functional annotations and generated estimates for stratified genetic variance 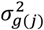 and stratified heritability *h*^2^ _*j*_ for each annotation. The heritability enrichment in each annotation was defined as 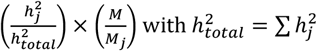 across all annotations.

When covariates were fitted as random effects, we model heteroscedasticity in error terms ***e***, i.e, 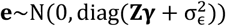, where **Z** is a *N* × *P* matrix of covariates to be fitted as random effects, and **γ** is a *P*-dimensional vector of random effects, and 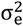 denotes the residual variance effect. Overall, the phenotypic variance can be expressed as

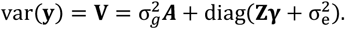

As 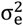 depends on the mean of each covariate in the **Z** matrix, we then assume the phenotypic variance to be one and define the heritability *h*^2^ as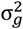.

The test statistics of heritability was calculated by Likelihood Ratio Test. The sampling variance of the variance component estimations were calculated by the diagonal elements of inverse of the Fisher information matrix. Permutation tests were performed by randomly shuffling the phenotypes across individuals. The permutation p-value was determined by the number of permutations with estimates exceeding the point estimate in the original model divided by the number of permutations (10,000 in this study).

### Simulation for PS

We simulated a phenotype (*y*) impacted by a localized environmental effect based on real birth region information in UKB. The model is

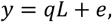

where *L* is a standardized indicator variable (mean 0, variance 1) for one specific region, 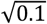 is the effect size, *e*∼*N*(0,0.9) is the error term. This setup ensures that the phenotype has a mean of 0 and a variance of 1, with the selected region explaining 10% of the total phenotypic variance.

We selected two birth regions—London and Sheffield—based on their contrasting average SC values, with London among the highest and Sheffield among the lowest (see **Supplementary Table 1**). We consider it unrealistic for a localized environmental effect to explain more than 10% of phenotypic variance. For context, a region comprising approximately 2% of the total sample (2.65% for Sheffield and 1.35% for London) would require an effect size of 2.26 standard deviations to explain 10% of the variance. This would correspond to an average difference of 20.9 cm in height, 2.78 children for NC, and 12.5 years of education—magnitudes that are implausibly large for regional effects.

### Simulation for scale transformation

We simulated a phenotype (*y*) where singletons have a directional effect based on real singleton genotype in UKB. The model is

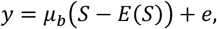

where *µ*_*b*_ is the directional effect size, ranging from -0.01 to +0.01 in increments of 0.005, *S* is the SC, *E*(*S*) = 17.43 is the mean SC, 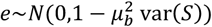 is the error term, and var(*S*) = 124.11 is the variance of SC.

To simulate a scale transformation effect, we mapped the simulated phenotype (as defined in the equation above) from its underlying Gaussian scale to an observed scale, based on the empirical distribution of a real trait in UKB.

Assume a real trait has *K* unique values *x*_1_, *x*_2_, …, *x*_*K*_ (ordered from smallest to largest), with corresponding proportions *n*_1_, *n*_2_, …, *n*_*K*_ . We first computed the cumulative proportions and used the standard normal quantile function to determine the corresponding cut points on the Gaussian scale. Each interval between two adjacent cut points was then mapped to one of the unique values *x*_1_, *x*_2_, …, *x*_*K*_, such that simulated phenotype values falling within a given interval were assigned the corresponding observed trait value.

### URLs

- PLINK2: https://www.cog-genomics.org/plink/2.0/
- GCTA: https://yanglab.westlake.edu.cn/software/gcta/#Overview
- Target regions for UKB WES releases: https://biobank.ndph.ox.ac.uk/ukb/ukb/auxdata/xgen_plus_spikein.GRCh38.bed
- <u>WES annotation helper files:</u> https://biobank.ctsu.ox.ac.uk/crystal/refer.cgi?id=916
- diagGREML R implementation: https://github.com/huanweiuq/diagGREML

## Data Availability

This study is based on data from the UK Biobank, access to which can be obtained by application.

https://www.ukbiobank.ac.uk/use-our-data/apply-for-access/

## ACKNOWLEDGEMENTS

We thank the participants of the UK Biobank for enabling this research. UKB data was accessed under approved project ID 12505. L.Y. is supported by the Snow Medical Research Foundation and the Australian Research Council (grant no. FT220100069). P.M.V. was funded by the Australian Research Council (grant no. FL180100072), the Australian National Health and Medical Research Council (grant no. 113400) and acknowledges funding from the European Research Council (grant 101198904). We also acknowledge support from the National Institutes of Health (grant no. NIH R01 MH100141) and the University of Queensland Foundation Research Excellence Award.

## AUTHOR CONTRIBUTIONS

L.Y. and P.M.V. designed the study. L.Y., P.M.V., and M.E.G. supervised the work. H.W. performed the analyses with the assistance from P.W., J.S, M.F., Y.Z., K.E.K., Z.Z., V.H., J.Z., H.W. and L.Y. wrote the paper with contributions from all authors.

## REFERENCES

1 Wainschtein, P. et al. Estimation and mapping of the missing heritability of human phenotypes. Nature (2025). 10.1038/s41586-025-09720-6

2 Steinsaltz, D., Dahl, A. & Wachter, K. W. On Negative Heritability and Negative Estimates of Heritability. Genetics 215, 343–357 (2020). 10.1534/genetics.120.303161

3 Rocheleau, G. et al. Rare variant contribution to the heritability of coronary artery disease. Nat Commun 15, 8741 (2024). 10.1038/s41467-024-52939-6

4 Taliun, D. et al. Sequencing of 53,831 diverse genomes from the NHLBI TOPMed Program. Nature 590, 290–299 (2021). 10.1038/s41586-021-03205-y

5 Hernandez, R. D. et al. Ultrarare variants drive substantial cis heritability of human gene expression. Nat Genet 51, 1349–1355 (2019). 10.1038/s41588-019-0487-7

6 Yang, J. et al. Common SNPs explain a large proportion of the heritability for human height. Nat Genet 42, 565–569 (2010). 10.1038/ng.608

7 Yang, J., Lee, S. H., Goddard, M. E. & Visscher, P. M. GCTA: a tool for genome-wide complex trait analysis. Am J Hum Genet 88, 76–82 (2011). 10.1016/j.ajhg.2010.11.011

8 Weiner, D. J. et al. Polygenic architecture of rare coding variation across 394,783 exomes. Nature 614, 492–499 (2023). 10.1038/s41586-022-05684-z

9 Young, A. I. et al. Relatedness disequilibrium regression estimates heritability without environmental bias. Nat Genet 50, 1304–1310 (2018). 10.1038/s41588-018-0178-9

10 Zhang, Y. et al. The contribution of gametic phase disequilibrium to the heritability of complex traits. Nat Genet 57, 1418–1425 (2025). 10.1038/s41588-025-02192-4

11 Mathieson, I. & McVean, G. Differential confounding of rare and common variants in spatially structured populations. Nat Genet 44, 243–246 (2012). 10.1038/ng.1074

12 Zaidi, A. A. & Mathieson, I. Demographic history mediates the effect of stratification on polygenic scores. Elife 9 (2020). 10.7554/eLife.61548

13 Abdellaoui, A. et al. Genetic correlates of social stratification in Great Britain. Nat Hum Behav 3, 1332–1342 (2019). 10.1038/s41562-019-0757-5

14 1000 Genomes Project Consortium et al. A global reference for human genetic variation. Nature 526, 68–74 (2015). 10.1038/nature15393

15 Field, Y. et al. Detection of human adaptation during the past 2000 years. Science 354, 760–764 (2016). 10.1126/science.aag0776

16 Zhou, W. et al. SAIGE-GENE+ improves the efficiency and accuracy of set-based rare variant association tests. Nat Genet 54, 1466–1469 (2022). 10.1038/s41588-022-01178-w

17 Smyth, G. K. Generalized linear models with varying dispersion. Journal of the Royal Statistical Society: Series B (Methodological) 51, 47–60 (1989).

18 Lee, Y. & Nelder, J. A. Hierarchical generalized linear models. Journal of the Royal Statistical Society: Series B (Methodological) 58, 619–656 (1996).

19 Rönnegård, L., Shen, X. & Alam, M. hglm: A package for fitting hierarchical generalized linear models. The R Journal 2, 20–28 (2010).

20 Young, A. I., Wauthier, F. L. & Donnelly, P. Identifying loci affecting trait variability and detecting interactions in genome-wide association studies. Nat Genet 50, 1608–1614 (2018). 10.1038/s41588-018-0225-6

21 Falconer, D. S. The inheritance of liability to certain diseases, estimated from the incidence among relatives. Annals of human genetics 29, 51–76 (1965). 10.1111/j.1469-1809.1965.tb00500.x

22 Bycroft, C. et al. The UK Biobank resource with deep phenotyping and genomic data. Nature 562, 203–209 (2018). 10.1038/s41586-018-0579-z

23 Backman, J. D. et al. Exome sequencing and analysis of 454,787 UK Biobank participants. Nature 599, 628–634 (2021). 10.1038/s41586-021-04103-z

24 U. K. Biobank Whole-Genome Sequencing Consortium. Whole-genome sequencing of 490,640 UK Biobank participants. Nature 645, 692–701 (2025). 10.1038/s41586-025-09272-9

25 Wang, Y. et al. Theoretical and empirical quantification of the accuracy of polygenic scores in ancestry divergent populations. Nat Commun 11, 3865 (2020). 10.1038/s41467-020-17719-y

26 Chang, C. C. et al. Second-generation PLINK: rising to the challenge of larger and richer datasets. Gigascience 4, 7 (2015). 10.1186/s13742-015-0047-8

27 Gilmour, A. R., Thompson, R. & Cullis, B. R. Average information REML: an efficient algorithm for variance parameter estimation in linear mixed models. Biometrics, 1440–1450 (1995).

