## supplementary materials for "Pitfalls in estimating and interpreting the contribution of ultra-rare genetic variants to the heritability of complex traits"

Wang H. et al.

### SUPPLEMENTARY NOTES

#### Supplementary Note 1. Expectation of the bias of singleton-based heritability due to population stratification (Proof of Equation (2))

Consider a phenotype with a 2-group population structure explaining $q^2$ of phenotypic variance,

$$y = qL + e,$$

where

- $y$ is the phenotype
- $L$ is a standardized indictor variable for the population structure with mean 0 and variance 1, i.e. $L = \frac{l-\pi}{\sqrt{\pi(1-\pi)}}$ with $l$ the zero-one indictor variable and $\pi$ the proportion of ones
- $q$ is the effect size in the standardized unit, so the phenotypic variance explained by $L$ is $q^2$.
- $e$ is the error term from $N(0,1-q^2)$, so the phenotype $y$ has a mean 0 and variance 1

We consider the distribution of SC ($S$) differ in two population groups, with $S_0$and $S_1$ being the expectation of SC in the group of zeros and ones, respectively.

| $l$ | proportion | $L$ | $L^2$ | $E(S\|l)$ |
|---|---|---|---|---|
| 0 | 1-$\pi$ | $\frac{-\pi}{\sqrt{\pi(1-\pi)}}$ | $\frac{\pi}{1-\pi}$ | $S_0$ |
| 1 | $\pi$ | $\frac{1-\pi}{\sqrt{\pi(1-\pi)}}$ | $\frac{1-\pi}{\pi}$ | $S_1$ |

Then in the whole sample, the expectation of SC is

$$E(S) = E(S|l=0)\times(1-\pi) + E(S|l=1)\times\pi = S_0(1-\pi) + S_1\pi$$

The HE regression-based heritability estimate is essentially a simple linear regression between $y^2$ and $G$ with $G$ being $S/E[S]$.

$$\mathrm{E}\left[\widehat{h^2_{HE}}\right] = \frac{\mathrm{cov}(y^2, G)}{\mathrm{var}(G)}$$

The covariance between $y^2$ and $G$ can be expressed as

$$\begin{aligned}
\mathrm{cov}(y^2,G) &= \mathrm{cov}(q^2L^2 + 2qLe + e^2, G) = q^2\mathrm{cov}(L^2,G) + 2q\mathrm{cov}(Le,G) + \mathrm{cov}(e^2,G) \\
&= q^2\mathrm{cov}(L^2,G) + 2q\times 0 + 0 \\
&= q^2[E(L^2G) - E(L^2)E(G)] \\
&= q^2[E(L^2G|l=0)\times(1-\pi) + E(L^2G|l=1)\times\pi - 1\times 1] \\
&= q^2\left[\frac{\pi}{1-\pi}\frac{S_0}{E(S)}(1-\pi) + \frac{1-\pi}{\pi}\frac{S_1}{E(S)}\pi - 1\right] \\
&= \frac{(S_1-S_0)(1-2\pi)q^2}{S_0(1-\pi)+S_1\pi}
\end{aligned}$$

Therefore,

$$\mathrm{E}\left[\widehat{h^2_{HE}}\right] = q^2\times(1-2\pi)\times\frac{(S_1-S_0)}{S_0(1-\pi)+S_1\pi}\times\frac{1}{\mathrm{var}(G)} = q^2\Delta_S(1-2\pi)\left[\frac{E(S)}{\mathrm{var}(S)}\right]$$

where $\Delta_S = S_1 - S_0$.

#### Supplementary Note 2. The case of phenotypic variance difference between sub-populations

We consider the following model: $y = zy_1 + (1 - z)y_0$, where $z$ is the indicator of sub-population 1, $y_1$ and $y_0$ follow the same phenotypic distribution as individuals in population 1 and 0, respectively. We have that $\mathrm{E}[z] = \pi$ and denote $\mathrm{var}[y_k] = \sigma_k^2$ and $\lambda = \sigma_1^2/\sigma_0^2$.

For simplicity, we assume that $\mathrm{E}[y_0] = \mathrm{E}[y_1] = 0$ and $\mathrm{var}[y] = \pi\sigma_1^2 + (1 - \pi)\sigma_0^2 = 1$.

Therefore, $\sigma_0^2 = 1/[\pi(\lambda - 1) + 1]$ and $\sigma_1^2 = \lambda/[\pi(\lambda - 1) + 1]$

and $S_0 = \mathrm{E}[\mathrm{S}] - \pi\Delta_S$ and $S_1 = \mathrm{E}[\mathrm{S}] + (1 - \pi)\Delta_S$

$$
\begin{aligned}
\mathrm{cov}(\mathrm{S}, \mathrm{y}^2) &= \mathrm{E}[\mathrm{Sy}^2] - \mathrm{E}[\mathrm{S}]\mathrm{E}[\mathrm{y}^2] \\
&= \pi S_1\sigma_1^2 + (1 - \pi)S_0\sigma_0^2 - \mathrm{E}[\mathrm{S}] \\
&= \sigma_0^2[\pi S_1(\lambda - 1) + \mathrm{E}[\mathrm{S}]] - \mathrm{E}[\mathrm{S}] \\
&= \frac{\pi(\lambda - 1)[\mathrm{E}[\mathrm{S}] + (1 - \pi)\Delta_S] + \mathrm{E}[\mathrm{S}] - [\pi(\lambda - 1) + 1]\mathrm{E}[\mathrm{S}]}{\pi(\lambda - 1) + 1} \\
&= \frac{\pi(1 - \pi)\Delta_S(\lambda - 1)}{\pi(\lambda - 1) + 1}
\end{aligned}
$$

Therefore,

$$
\mathrm{E}\left[\widehat{h_{SC}^2} \middle| h_{SC}^2 = 0\right] = \frac{\mathrm{cov}(\mathrm{G}, \mathrm{y}^2)}{\mathrm{var}(G)} = \pi(1 - \pi)\Delta_S \left(\frac{\lambda - 1}{\pi(\lambda - 1) + 1}\right)\left[\frac{E(S)}{\mathrm{var}(S)}\right]
$$

In the absence of variance difference between sub-populations, $\lambda = 1$ and $\mathrm{E}\left[\widehat{h_{SC}^2} \middle| h_{SC}^2 = 0\right] = 0$.

#### Supplementary Note 3. Empirical distribution of SC in a European ancestry population

We analysed whole-exome sequencing (WES) data from 469,835 UKB participants, from which we selected a subset of 305,813 unrelated individuals with European ancestries (METHODS). Among the 12,894,284 biallelic autosomal sequenced SNPs, 5,330,210 (41.3%) were singletons, including 2,723,052 located within coding genomic regions (**URLs**). On average, each individual included in our study sample carried ~17 singletons. The SD of SC is ~11. The distribution of SC (**Supplementary Fig. 3a**) showed a strong over-dispersion (range: 0 – 356).

Overdispersion in SC distribution reflects widespread positive covariances between singletons across the genome, that is induced by a combination of within-chromosome linkage disequilibrium (LD) between singletons and population structure. Given that ~95% of these pairwise covariances involve singletons located on different chromosomes, the observed over-dispersion can largely be attributed to population structure.

We found significant correlations between SC and several population structure-related variables, including common-variants based principal components (PCs), 22 assessment centres (AssCen), birth place coordinates (BCs), 378 local birth regions (BRs), the proportion of non-European local ancestries as determined by RFMix[28], and the percentage of migration singletons (**Supplementary Fig**. 3). In addition, SC was correlated with demographic variables such as year of birth (YOB) and relatedness, likely reflecting ascertainment of study participants or recent migration. We found no significant correlations between SC and sex as well as between SC sequencing depth

The odd- and even-chromosome SC correlation, an indicator for population structure, was found to be high both in the full sample (r=0.723) and in each local BR (**Supplementary Fig 4**), while it could be attenuated by fitting some population covariates jointly (**Supplementary Fig 4**). We also conducted association study of HapMap 3 (HM3) SNPs on even chromosomes with SC on odd chromosomes, which shown widespread inflation, which could be attenuated by adjusting common PCs (**Supplementary Fig 5**). Altogether, these results indicated the existence of uncaptured population structure.

#### Supplementary Note 4. The impact of truncation in a true model

Image a true model $y_i \sim N(0, \frac{h^2 N S_i}{M} + (1 - h^2))$, where $h^2$ is the heritability, N is the sample size, M is the number of singletons, and $S_i$ is the singleton count for i-th individual in the whole sample.

Let $\sigma_i$ be the singleton count for i-th individual in the truncated sample. The expected value of heritability estimates in the truncated sample will be

$$E(h_T^2) = \frac{\mathrm{cov}\left(\frac{NS_i}{M}, \frac{N_T \sigma_i}{M_T}\right)}{\mathrm{var}\left(\frac{N_T \sigma_i}{M_T}\right)} h^2 = \frac{E(\sigma_i)\mathrm{cov}(S_i, \sigma_i)}{E(S_i)\mathrm{var}(\sigma_i)} h^2,$$

where $N_T$ and $M_T$ are the sample size and the number of singletons in the truncated sample. In our UKB data, $E(\sigma_i)$ is 16.6, $E(S_i)$ is 17.4, $\mathrm{cov}(S_i, \sigma_i)$ is 32.9, and $\mathrm{var}(\sigma_i)$ is 36.9. So, the $E(h_T^2)$ is $0.85h^2$

### SUPPLEMENTARY FIGURES

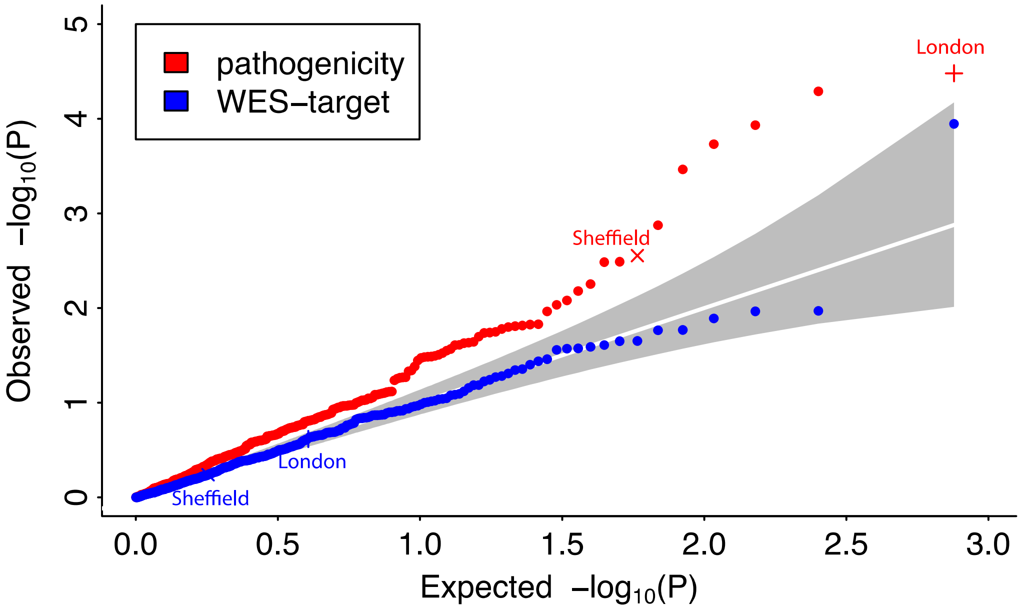

Supplementary Figure 1. The QQ-plot for 378 chi-square tests between each birth region (BR) with pathogenicity or WES-target annotations. The inflation factors (lambda_GC) are 1.637 for pathogenicity annotation and 0.993 for WES-target annotation. Each dot represents a BR. The pathogenicity functional annotations and WES target regions were downloaded from UKB (**URLs**; Methods). Pathogenicity functional annotations were derived from snpEff. Singletons annotated as "synonymous" or lacking annotation information were labelled as "neutral". The joint distribution of SC in three pathogenicity categories and BRs can be found in Supplementary Table 2.

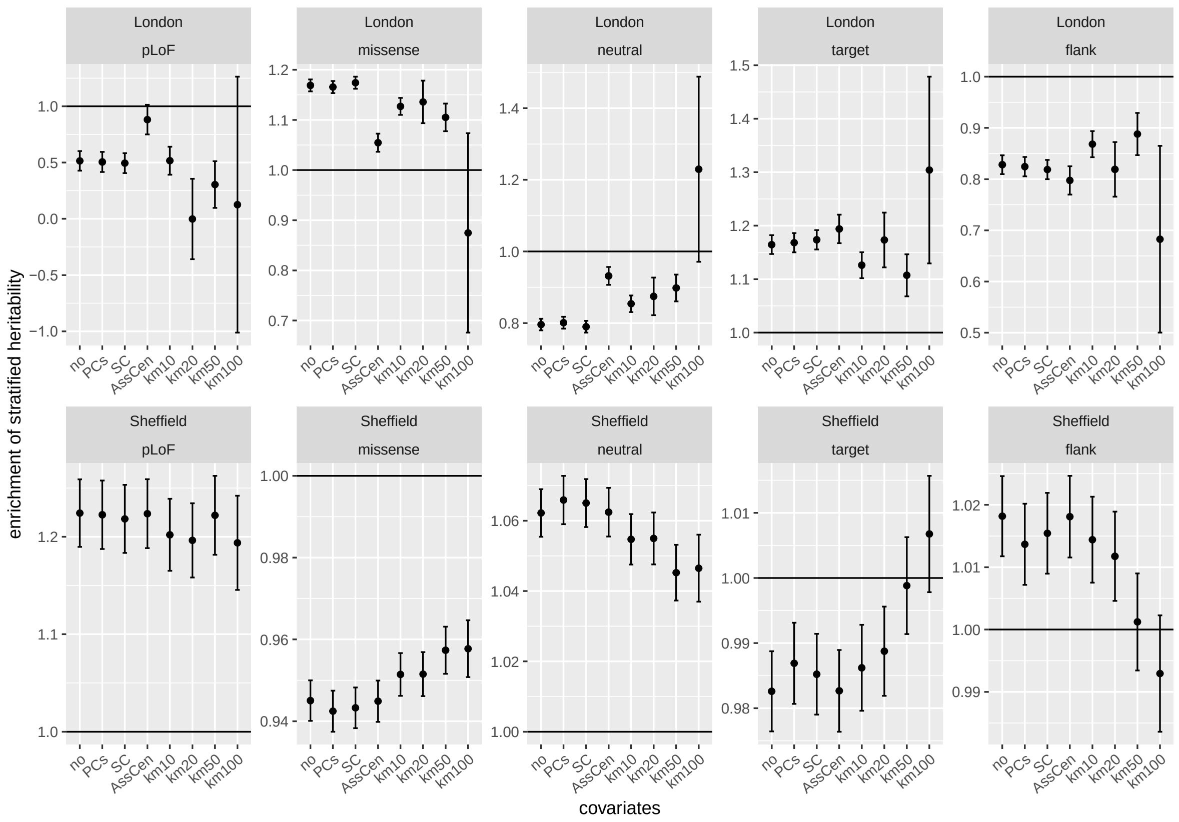

**Supplementary Figure 2. The per-SNP $\widehat{h^2_{SC}}$ enrichments estimated by the GREML models for pathogenicity and WES-target annotations in the PS-based simulation model.** We fitted covariates individually in the fixed component of the GREML model, including principal components (PCs), singleton count (SC), assessment centres (AssCen), and k-means BC clustering with 10, 20, 50, and 100 clusters. Error bars represent 1.96 times standard errors.

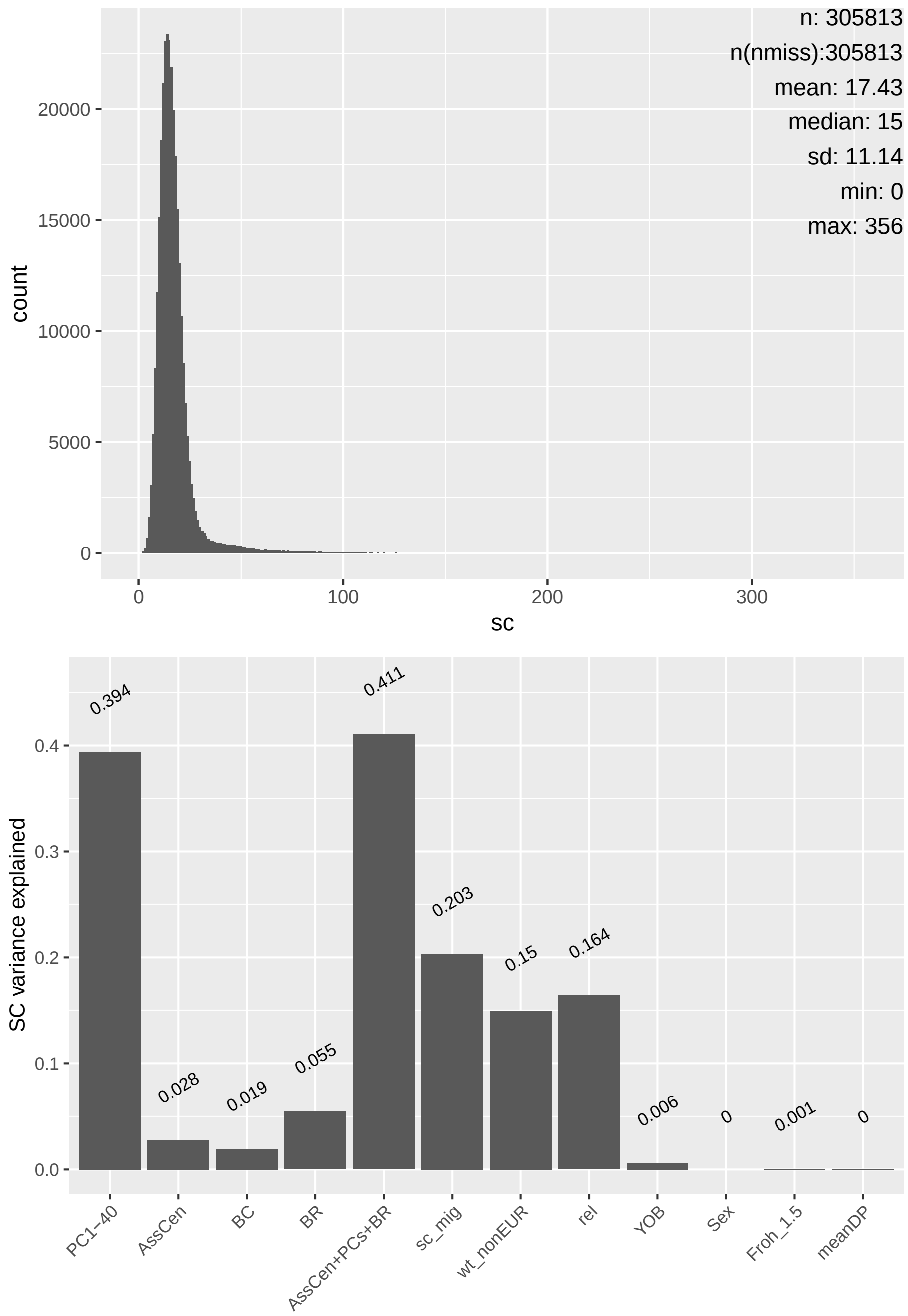

Supplementary Figure 3. Distribution and phenotypic correlates of singleton count (SC). The top panel represents the distribution of SC and the bottom panel shows the variance of SC explained by different sets of covariates. Birthplace coordinates (BCs) were recorded during the UKB assessment visits (Data Fields 129 and 130. Birth regions, derived from BCs, were defined based on UK local authorities division. Weighted estimated probability as non-European ancestry (wt_nonRUR) was generated by RFMix based on 1000 genome project excluding individuals from AMR population. Relatedness (rel) was measured by the sum of genetic relationship matrix (GRM) off-diagonal elements based on common (MAF>=0.05) Hapmap 3 genotyped and imputed SNPs among 305,813 individuals. The migration singletons (sc_mig) were defined that any singletons also present in any AFR, EAS, or SAS populations in 1000 genome project (phase 3) data. The inbreeding coefficient (Froh_1.5) indicated runs of homozygosity > 1.5 Mb. The mean sequence depth (meanDP) across variants per sample was based on WES 200k release.

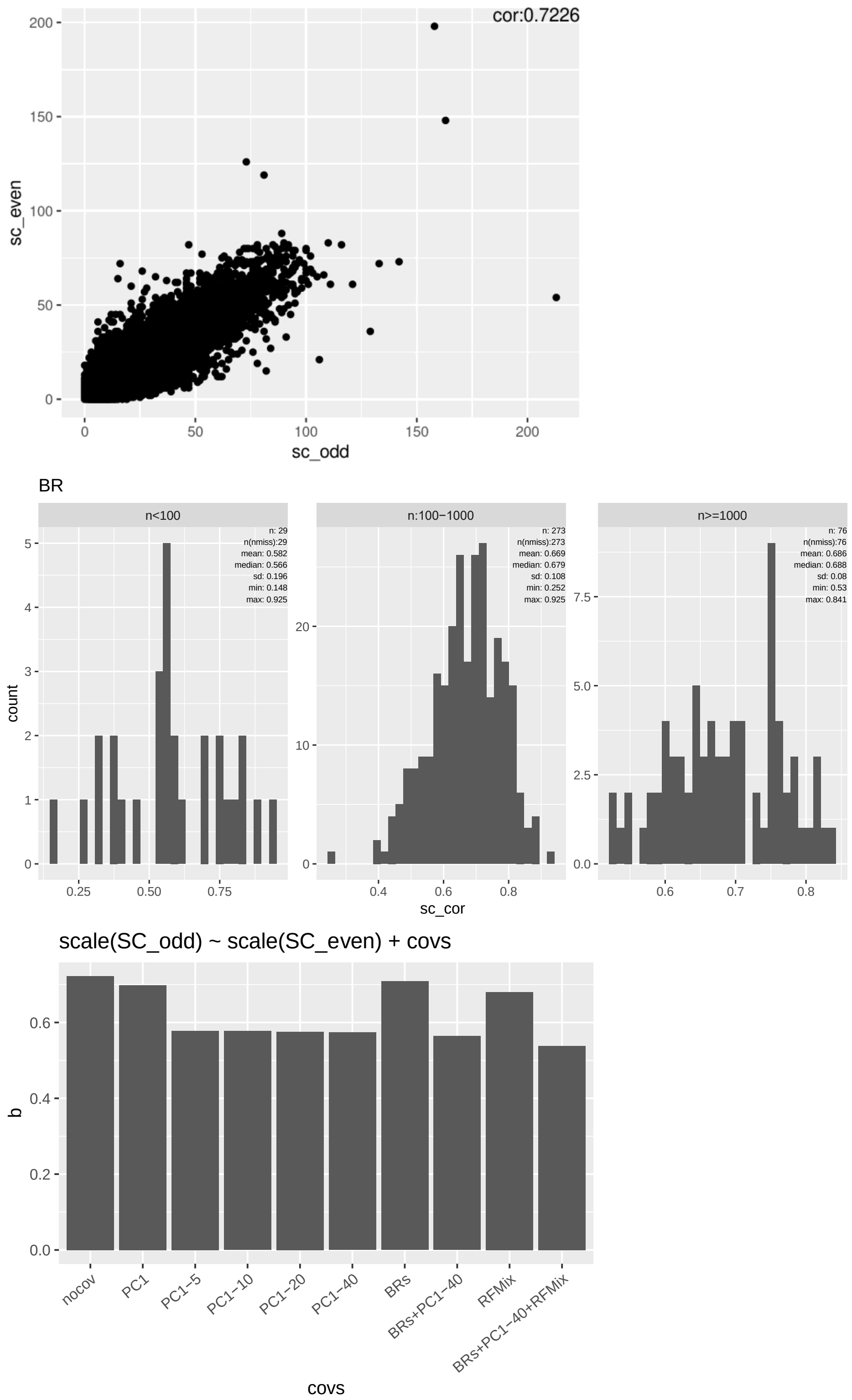

**Supplementary Figure 4. Odd- and even-chromosome SC correlation in the whole sample (a), each of 378 BR (b), and the whole sample corrected with different covariates (c).** The correlation between odd- and even-chromosome SC correlation was 0.72 in the whole sample. In panel b, we reported the distribution of odd- and even-chromosome SC correlations in each BR stratified by the sample size category of BRs. The covariate-adjusted correlation was calculated by standardizing (mean 0 and variance 1) the odd- and even-chromosome SC values and fitting covariates in a linear regression model.

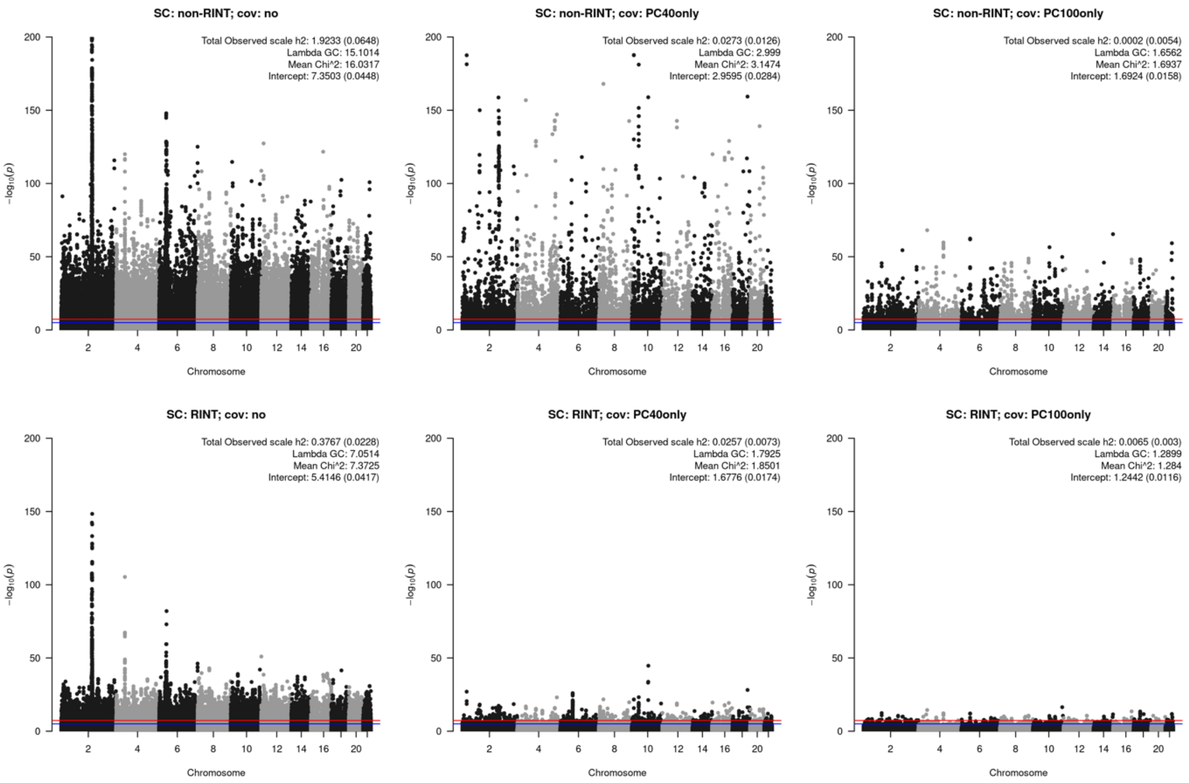

Supplementary Figure 5. Association study of even chromosome HM3 SNPs (m=633,888) with the odd chromosome SC. We applied or did not apply rank-based inverse normal transformation (RINT) and fitted different numbers of common PCs. Output from LD score regression analyses (observed scale heritability, lambda GC, mean chi-square, and intercept) are showed on the top right corner of each panel.

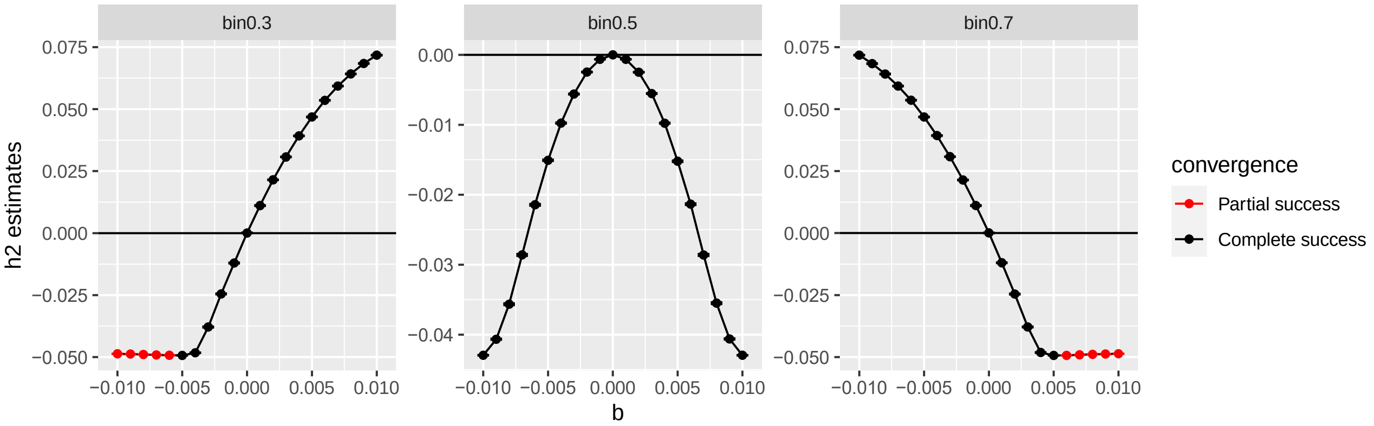

Supplementary Figure 6. The impact of binary scale transformation on the SC-based heritability in the presence of a direct mean effect. The simulated direct mean effects are shown on the x-axis. The heritability estimates on the y-axis were obtained from the GREML model with SC fitted as fixed effects. “binX” indicates that X proportion of the values were classified as 0s. "Partial success" refers to simulation situations where the GREML analysis failed to converge for some replicates. This occurred when the estimated values were expected to be negative with large magnitudes.

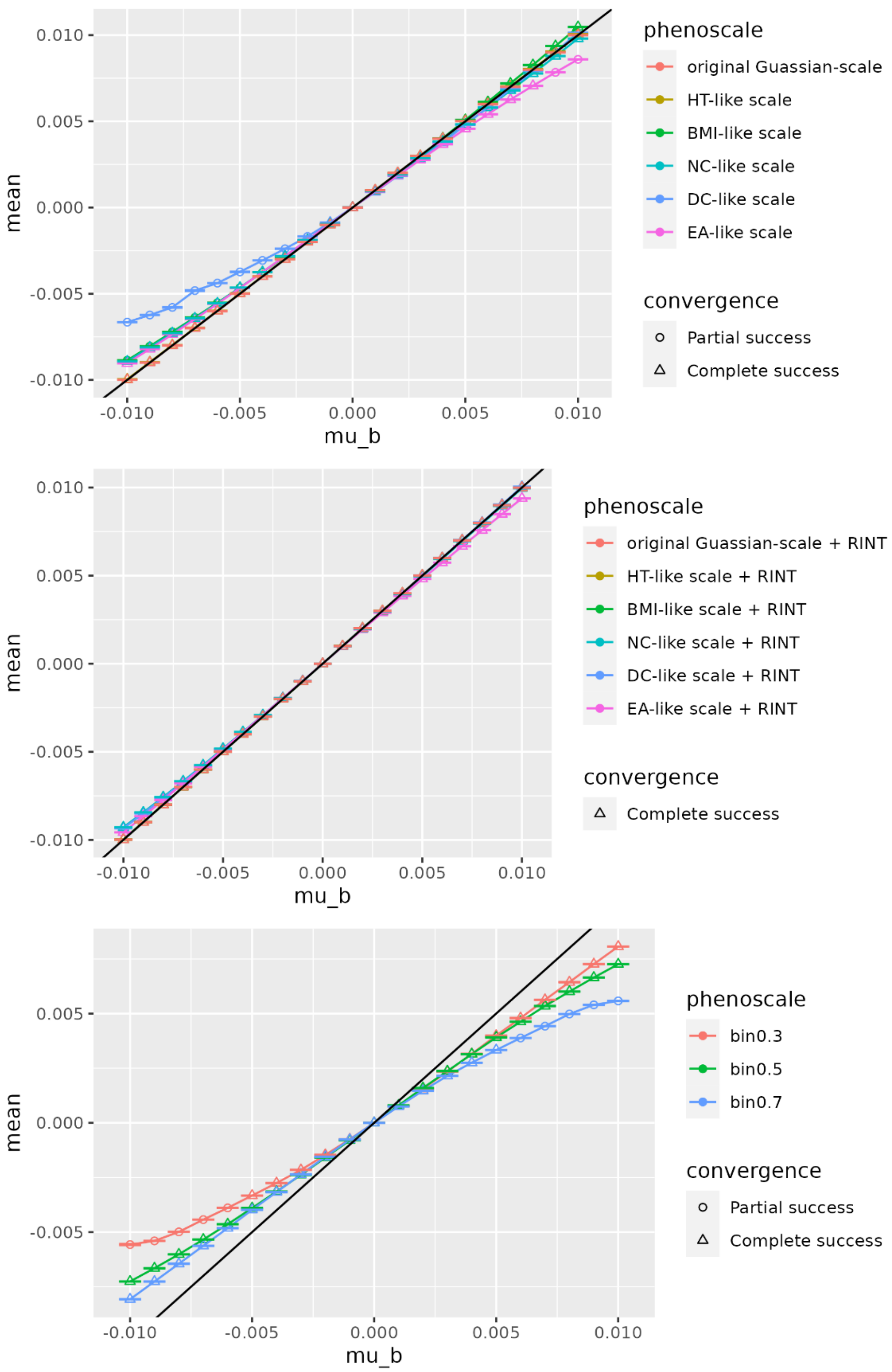

**Supplementary Figure 7. The impact of scale transformation on the mean effect estimates of SC on the phenotype.** The true values of the parameter $\mu_u$ were shown on the x-axis, while the averages of the mean effect estimates were plotted on the y-axis. The black line represents a reference line with slope 1 and intercept 0.

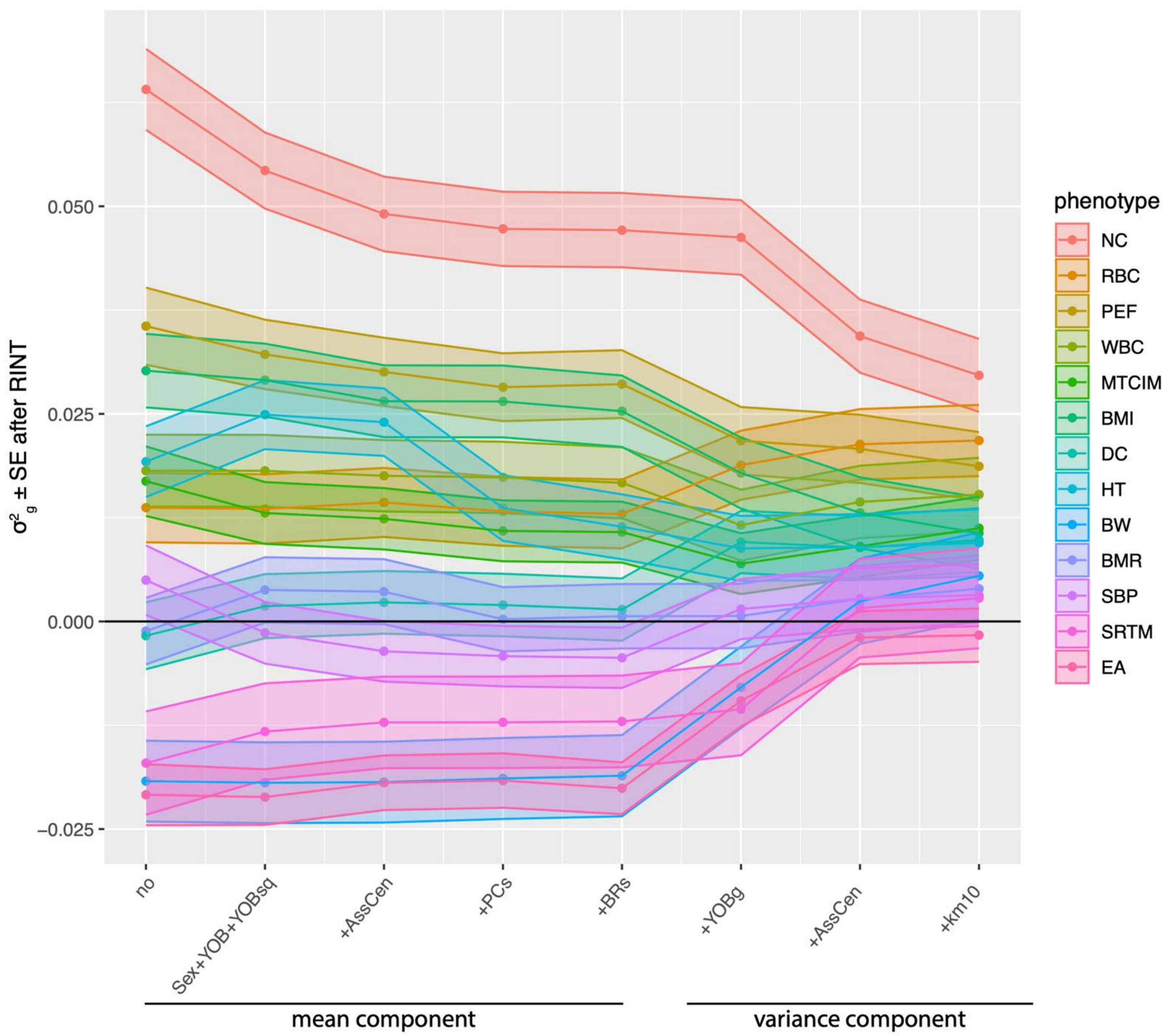

**Supplementary Figure 8. Fitting different covariates as the fixed or/and random effects for 13 traits after applying RINT to the phenotypes.** We sequentially fitted covariates as fixed effects from sex, year of birth (YOB), YOB squared (YOBsq), 22 assessment centres (AssCen), top 40 PCs, and 378 BRs, and then sequentially added covariates as random effects from seven YOB groups (YOBg), assessment centres, and 10 k-means BC clusters. Confidence bands represent standard errors from the GREML model.

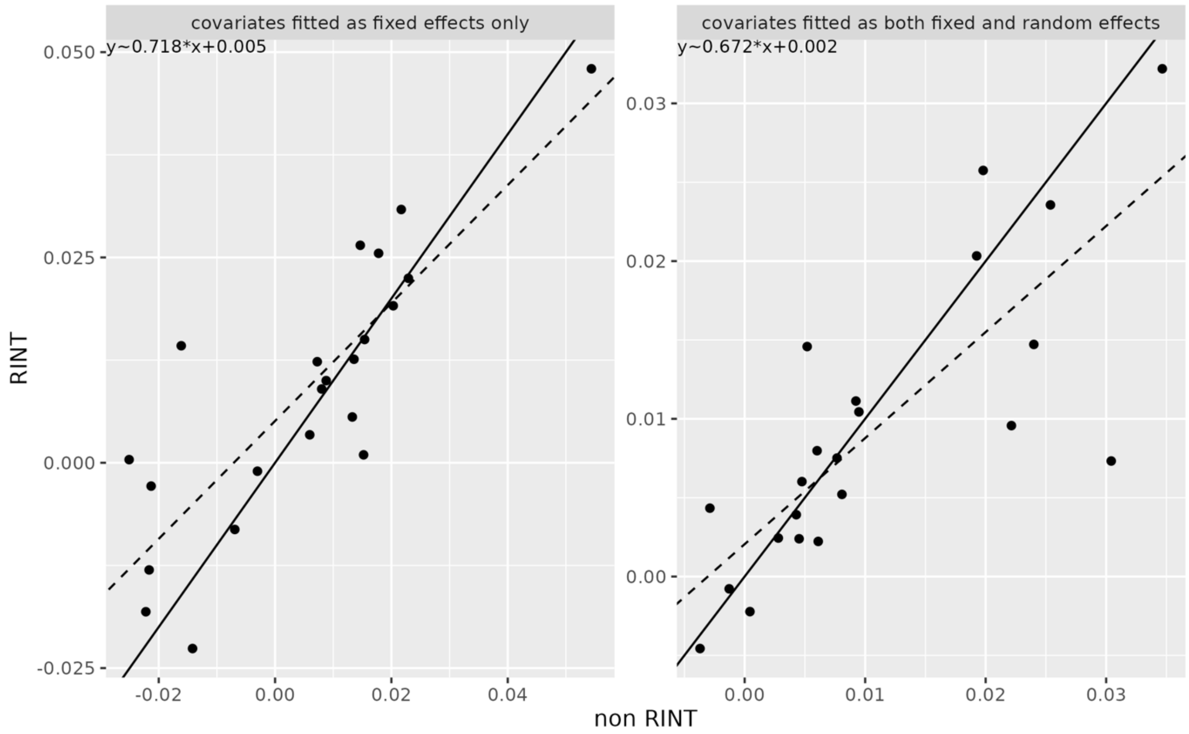

**Supplementary Figure 9. Effect of Rank-based Inverse Normal Transformation (RINT) on $\widehat{h^2_{SC}}$ estimates for 22 traits measured in the UK Biobank.** The x- and y-axis show estimates obtained from untransformed and RINT-transformed phenotypes, respectively. The left panel showed the results with covariates fitted as fixed effects only, and the right panel showed the results with covariates fitted as both fixed and random effects. Dash line is the regressed line and solid line is the line with slope 1 and intercept 0. Each dot represents a trait.

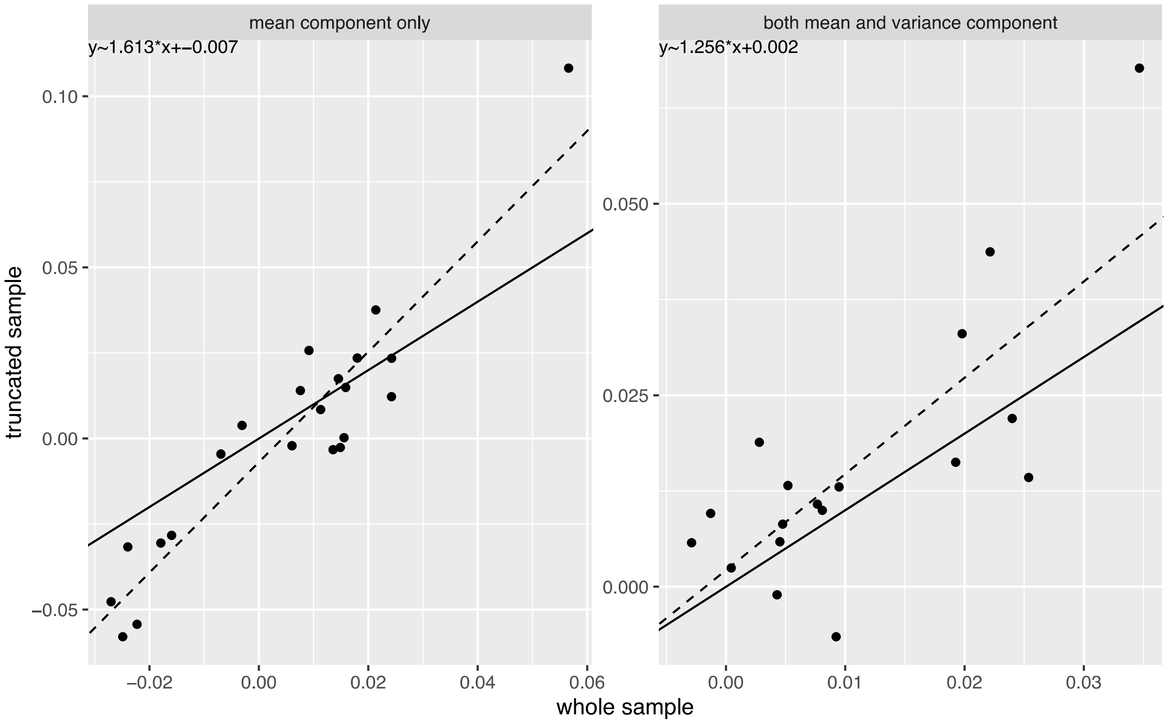

**Supplementary Figure 10. Compare $\widehat{h^2_{SC}}$ estimates in the whole sample or truncated sample across 22 traits.** The fitted liner regression equations were showed in the top left. The left panel showed the results with covariates fitted as fixed effects only, and the right panel showed the results with covariates fitted as both fixed and random effects. Dash line is the regression line and solid line is the line with slope 1 and intercept 0.
